# Comprehensive Evaluation of Human Donor Liver Viability with Polarization-Sensitive Optical Coherence Tomography

**DOI:** 10.1101/2025.03.31.25321497

**Authors:** Feng Yan, Qinghao Zhang, Bornface M. Mutembei, Chen Wang, Zaid A. Alhajeri, Kaustubh Pandit, Fan Zhang, Ke Zhang, Zhongxin Yu, Kar-Ming Fung, Shaima N. Elgenaid, Paige Parrack, Walid Ali, Clint A. Hostetler, Ashley N. Milam, Bradon Nave, Ron Squires, Paulo N. Martins, Narendra R. Battula, Steven Potter, Chongle Pan, Yu Chen, Qinggong Tang

## Abstract

Human liver transplantation is severely constrained by a critical shortage of donor livers, with approximately one quarter of patients on the waiting list dying due to the scarcity of viable organs. Current liver viability assessments, which rely on invasive pathological methods, are hampered by limited sampling from biopsies, particularly in marginal livers from extended criteria donors (ECD) intended to expand the donor pool. Consequently, there is a pressing need for more comprehensive and non-invasive evaluation techniques to meet the escalating demand for liver transplants. In this study, we propose the use of polarization-sensitive optical coherence tomography (PS-OCT) to perform a thorough viability evaluation across the entire surface of donor livers. PS-OCT imaging was conducted on multiple regions, achieving near-complete coverage of the liver surface, and the findings were cross-validated with histopathological evaluations. The analysis of hepatic parameters derived from pathology highlighted tissue heterogeneity. Leveraging machine learning and texture analysis, we quantified hepatic steatosis, fibrosis, inflammation, and necrosis, and established strong correlations (≥ 80%) between PS-OCT quantifications and pathological assessments. PS-OCT offers a non-invasive assessment of liver viability by quantifying hepatic parenchymal parameters across the entire donor liver, significantly complementing current pathological analysis. These results suggest that PS-OCT provides a robust, non-invasive approach to assessing donor liver viability, which could potentially decrease the discard rate of higher risk livers, thereby expanding the donor pool and reducing the inadvertent use of those livers unsuitable for transplantation.

## INTRODUCTION

Liver transplantation (LT) is the only viable therapy for patients experiencing extremely severe liver diseases such as cirrhosis and carcinomas at an advanced stage (*1, 2*). The demand for liver transplants has grown significantly in recent decades (*3*), and more than 10,000 candidates were waiting for liver allografts in 2023 in the US (with over 10,000 performed liver transplants) (*4*). A shortage of donor livers remains a major limitation and accounts for a large proportion of waitlist mortality (*5*). With the increasing number of waitlisted candidates and the rising number of patients dying while waiting (1 of 4 patients died on the liver waiting list in the US in 2023 (*6*)), efforts have been made in recent decades to increase the utilization of so-called marginal donor livers; such as livers from donors with advanced age, steatosis, ischemia, and hepatitis C virus-positive infecti(*7–10*). However, the current standard methods for screening marginal livers fail to provide transplant surgeons with sufficient information to make informed decisions, leading to a discard rate of nearly 70-80% for these types of donor livers (*11, 12*). Liver donor risk index (LDRI) is used to inform the viability of donor livers via evaluating independent donor characteristics such as age, cause of death, and split/partial allografts, but over 70% of physicians believe LDRI inadequately predicts the risk of organ failure or primary nonfunction, leading to persistently high discard rates (*13, 14*).

Pretransplant liver biopsy is the current gold standard for assessing these conditions in donor livers (*15, 16*). Hepatic steatosis, fibrosis, inflammation, and necrosis are four key parameters for evaluating donor liver viability from the biopsied tissue, with steatosis being the most dominant. Steatosis, often induced by nonalcoholic fatty liver disease, is highly prevalent in donor livers and can impact transplantation outcomes by increasing cold ischemic injury and impairing hepatic regeneration (*16–20*). It is associated with early allograft dysfunction, primary nonfunction, and postreperfusion syndrome (*15, 16, 21–23*). Additionally, elevated levels of fibrosis, inflammation, and necrosis correlate with increased graft dysfunction, reduced bile production, and impaired metabolic function, impacting both pre-transplantation evaluation and post-transplantation outcomes (*15, 24–27*).

For the biopsied liver tissue, Hematoxylin & Eosin (H&E) staining is routinely performed for histopathological assessment of liver quality. Additionally, stains like Periodic acid–Schiff (PAS), Trichrome, Periodic acid–Schiff–diastase (PASD), and Prussian Blue Iron are essential for evaluating steatosis, fibrosis, inflammation, and necrosis in donor livers (*28–30*). To minimize damage to the liver, only one or two tissue samples are taken from the liver during the biopsy in current clinical practice. Therefore, liver biopsy is invasive but suffers from a significant risk of sampling errors, limiting the utility of biopsy (*31, 32*). Liver biopsy analysis evaluates only one or 2 tiny fractions of the organ, providing an incomplete assessment of liver viability that does not adequately address the concerns transplant surgeons have about the condition of organs, especially those from deceased donors. An even worse scenario is that some high-risk livers get transplanted due to favorable biopsy findings in the tiny fraction of the liver evaluated. Therefore, there is a critical need for a technique that can achieve noninvasive evaluation of the entire liver to help determine graft viability.

Optical coherence tomography (OCT) is a noninvasive, label-free, high-speed imaging technique for depth-resolved imaging in real time (*33*). At present, OCT is widely employed for providing structural imaging in diverse domains, encompassing the examination of the human retina, cancerous tissues, various organs, vasculature, and applications within the field of dentistry (*34–41*). Specifically, OCT has demonstrated significant potential in characterizing microstructures of human organs, including kidney tubules and glomeruli (*36, 42*), as well as liver portal tracts (*37*). Moreover, OCT has undergone extensive development to yield enhanced tissue and functional information, including the imaging of fibrosis or collagen through polarization-sensitive OCT (PS-OCT) (*43, 44*). Despite these advances, no OCT studies have yet been conducted to establish quantitative assessment criteria for the viability of pre-transplant livers. The integration of structural and functional information through PS-OCT provides a novel approach to bridging the gap between research and clinical application in the pre-transplantation assessment of livers.

In this study, we first quantitatively evaluated the heterogeneity of hepatic tissue on deceased donor livers. After that, we systematically investigated the potential of PS-OCT for comprehensive imaging of deceased donor livers in a non-invasive and near real-time fashion. We utilized PS-OCT intensity and polarization images to correlate with liver biopsies in hepatic steatosis, fibrosis, inflammation, and necrosis with the assistance of structural tissue features and machine learning to build evaluation criteria for donor livers. After correlating assessment parameters and establishing screening thresholds for pathologically confirmed cases, we conducted a double-blind cross-test to evaluate the accuracy of PS-OCT. Our results introduce a noninvasive imaging method for generating a near real-time, color-coded viability map of the entire donor liver, thereby offering a comprehensive visualization and evaluation method to determine organ suitability for transplantation. This work highlights the translational potential and feasibility of PS-OCT in pre-transplantation donor liver evaluation.

## RESULTS

### Heterogeneous Distribution of Hepatic Tissues on Deceased Donor Livers

We systematically mapped the distribution patterns of four key clinical histopathologic parameters - steatosis, fibrosis, inflammation, and necrosis across the entire donor liver. The liver was sectioned into 20-40 regions to comprehensively cover its surface. Fig. 1 illustrates the heterogeneous distribution of these parameters on the liver surface. Score mapping was performed over the entire liver surface using point Gaussian diffusion filtering, overlaying the scores on a virtual 3D liver model. The detailed reconstruction process of this virtual 3D score mapping is described in Fig. S1. The quantification of the histopathologic parameters and the assessment of donor liver suitability were based on selected sectioning regions. A widely adopted clinical standard for determining the low-risk or high-risk of donor livers based on pathology (Table 1) was applied to map the distribution of hepatic tissues throughout the donor liver (*15, 45, 46*).

**Fig. 1.**
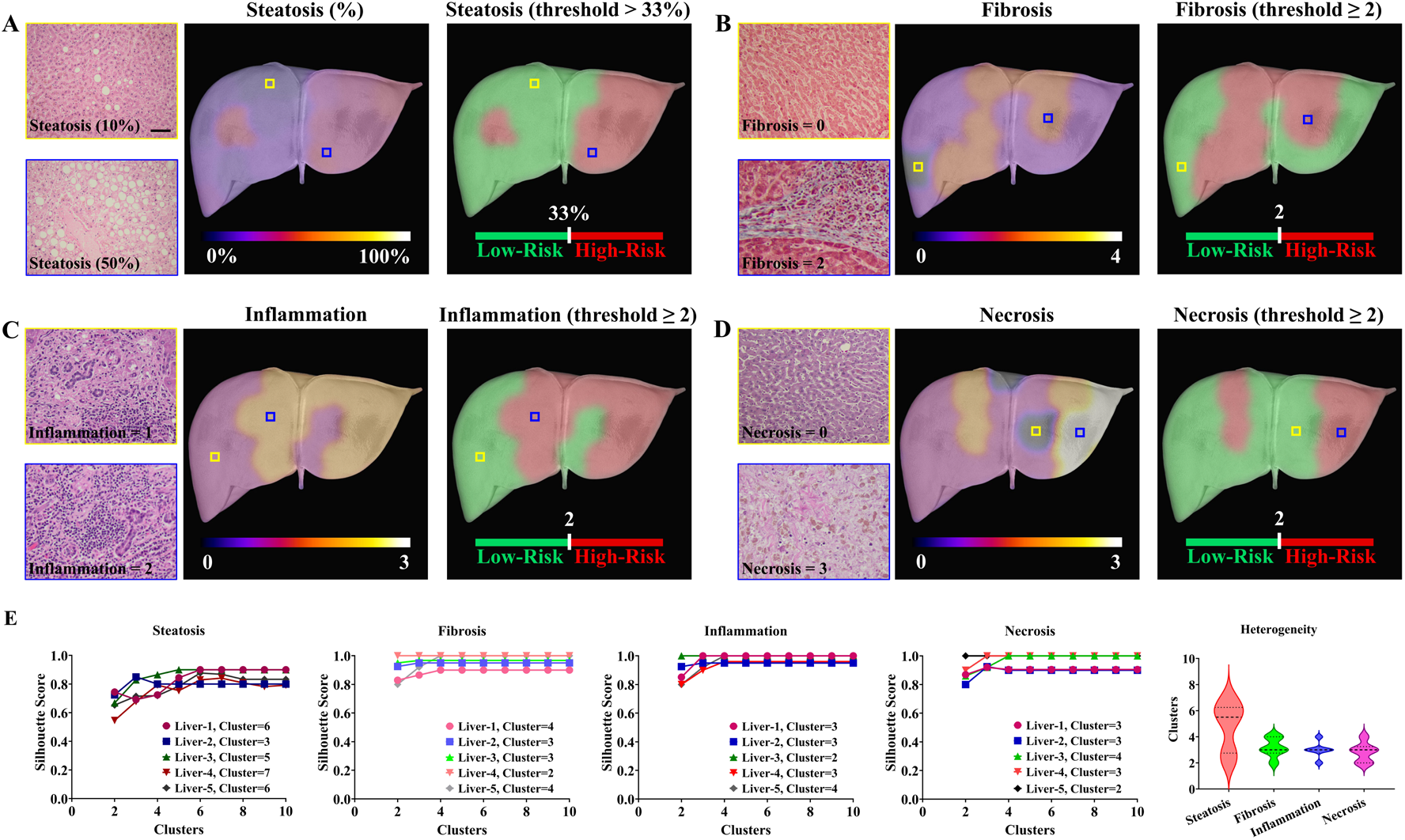
Distribution characters of hepatic histopathology on the entire donor liver. (A) Total steatosis (large and small droplets) and H&E stains. (B) Fibrosis and Trichrome stains. (C) Portal tract inflammation and PAS stains. (D) Hepatocellular or acidophilic necrosis and Iron stains. (E) Cluster statistics of hepatic parameters. The thresholds of steatosis, fibrosis, inflammation, and necrosis are 33%, 2, 2, and 2 from current clinical standards, respectively. The histopathologic distribution of these hepatic parameters is provided by 12 donor livers (n = 12).

**Table 1.** Selected Low-risk and High-risk Criteria for Liver Transplantation Based on Clinical Frozen Section Pathology. (***15, 45, 46***).

| Parameter | Low-Risk Criteria | High-Risk Criteria |
| --- | --- | --- |
| **Steatosis** (*macrovesicular steatosis*) | $\leq 33\%$ hepatocytes involved (score 0, 1) | $> 33\%$ hepatocytes involved (score 2, 3) |
| **Fibrosis** | Stage $< 2$ (score 0, 1) | Stage $> 2$ (score 2, 3, 4) |
| **Portal Tract Inflammation** | ≤ Mild (score 0, 1) | $>$ Mild (score 2, 3) |
| **Necrosis** | $\leq 4$ necrotic foci (score 0, 1) | $> 4$ necrotic foci (score 2, 3) |

Our results demonstrated a high degree of regional heterogeneity in degrees of steatosis (Fig. 1A), fibrosis (Fig. 1B), and necrosis (Fig. 1D), with the coexistence of mild steatosis (10%), non-fibrous (0), and non-necrotic (0) tissues alongside regions exhibiting extreme steatosis (50%), fibrosis (score 3), and necrosis (score 3) within the same donor liver. Inflamed hepatic tissues showed a gradual gradient of heterogeneity (scores 1 and 2) throughout the liver. Quantitative evaluation also revealed significant regional variability in inflammation (Fig. 1C), a factor crucial for assessing donor liver viability according to current clinical standards. H&E, Trichrome, and PAS biopsies highlighted varying degrees of hepatic steatosis, fibrosis, inflammation, and necrosis within representative samples from the same donor liver.

In Fig. 1E, a K-means unsupervised learning model was used to evaluate the heterogeneity degree of these four pathological parameters through clustering and silhouette analysis. The optimal number of clusters was determined by the highest silhouette scores, with higher cluster numbers indicating greater heterogeneity. Steatosis, fibrosis, inflammation, and necrosis exhibited multiple cluster distributions across the liver surface in five representative livers, highlighting the heterogeneity of these pathologic parameters across the organ. Among these parameters, steatosis demonstrated the greatest heterogeneity across the liver surface, due to its substantially higher clustering compared to fibrosis, inflammation, and necrosis.

### Global Recognition and Quantification of Hepatic Steatosis via PS-OCT Intensity Imaging

PS-OCT provided intensity and polarization images (including phase retardation, optic axis, and DOPU) to qualitatively and quantitatively assess hepatic microstructures (Fig. S2A). Our results showed that PS-OCT intensity images revealed distinct microstructural characteristics across various sites, highlighting the heterogeneity of hepatic tissue microstructures over the liver surface (Fig. S2B). Notably, PS-OCT intensity images enabled the direct detection of regions of steatosis, and these findings were confirmed by histologic evaluation (Fig. 2A). Steatosis was automatically segmented and extracted from PS-OCT intensity images (Fig. 2B), with red and blue frames indicating the enlarged steatosis structures before and after segmentation, respectively. Fig. 2C and Movie S1 illustrate a 3D reconstruction of steatosis within hepatic tissues, providing insights into its spatial distribution and allowing quantification of volumetric density (green frames).

**Fig. 2.**
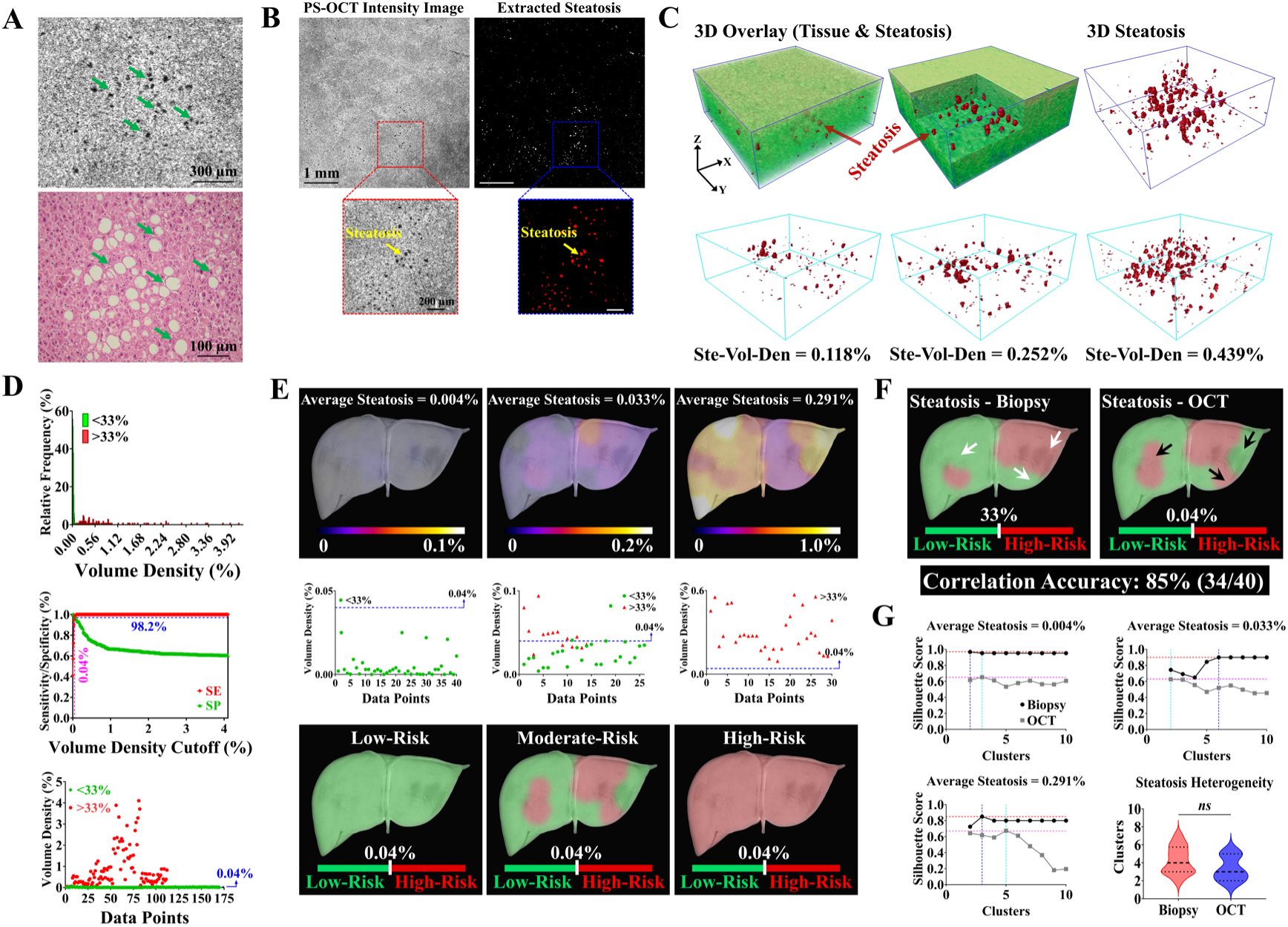
Segmentation and quantification of hepatic steatosis from PS-OCT intensity images for viability evaluation. (A) Steatosis recognition from PS-OCT intensity image and pathology. (B) Steatosis segmentation and extraction from PS-OCT images. (C) 3D reconstruction of steatosis for spatial visualization and quantification. (D) The extraction of steatosis density threshold for Low-Risk and High-Risk evaluations. (E) Comparison and distribution of steatosis over entire surfaces at low-risk, moderate-risk, and high-risk livers. Green spots indicate a pathologic steatosis score of <33% and red spots indicate a pathologic steatosis score of >33%. (F) Steatosis region comparison at Low-Risk and High-Risk between pathology and PS-OCT under clinical and distribution-based thresholds. (G) Cluster distribution comparison between pathology and PS-OCT. Ste-Vol-Den, steatosis volume density.

To establish a threshold for classifying steatosis density as low-risk or high-risk based on pathology, we plotted a histogram of steatosis volume density (%) (Fig. 2D). From the training dataset, an optimal threshold of 0.04% was identified, achieving a sensitivity of 0.98 and specificity of 0.97 in correlating PS-OCT evaluations with pathology. The scatter plot indicated that a steatosis density threshold of 0.04% effectively distinguished between low-risk (<33%) and high-risk (>33%) hepatic tissues. Fig. 2E presents three representative livers with low (0.004%), medium (0.033%), and high (0.291%) average steatosis densities, mapping the spatial distribution of steatosis over the liver surface. These PS-OCT images revealed a heterogeneous distribution of steatosis across the entire liver surface, validating concerns that tissue biopsy may be subject to significant sampling error. The scatter plots further classify steatosis density using the 0.04% PS-OCT threshold and compare it to pathology-based steatosis density.

For the liver with a low average steatosis density score, all scanned regions were classified as low-risk according to both the PS-OCT threshold of 0.04% and the pathology threshold of 33%, categorizing it as a low-risk donor liver. In contrast, the liver with a high average steatosis density score was uniformly classified as high-risk under both thresholds. The representative liver with a medium average steatosis density score contained both low-risk and high-risk regions based on PS-OCT and pathology thresholds, requiring further evaluation for a final risk assessment as moderate-risk. Using a steatosis density threshold of 0.04% for PS-OCT and 33% for pathology, we compared classifications across the liver surface, achieving a high correlation accuracy of 85% (34 out of 40 sites) (Fig. 2F).

In Fig. 2G, we observed different clustering distributions of steatosis density scores between PS-OCT and pathology. Donor livers classified as low-risk or high-risk had a higher number of clusters in PS-OCT (magenta + cyan lines) than in pathology (red + blue lines), while donor livers classified as moderate-risk showed fewer clusters in PS-OCT. Statistical analysis indicated that despite differences in clustering numbers, the variation between PS-OCT and pathology was not statistically significant.

### Extraction and Quantification of Hepatic Fibrosis via PS-OCT DOPU Imaging

We demonstrated that PS-OCT polarization images highlight the heterogeneous distribution of hepatic fibrosis across the entire liver (Fig. S2C). Hepatic fibrosis is graded on a clinical scale from 0 to 4 to evaluate donor liver viability. Fig. 3A provides pathology images of hepatic tissues at different fibrosis levels, along with representative 2D DOPU images, to illustrate the recognition of fibrosis at varying levels. We observed that PS-OCT DOPU images effectively visualized and classified fibrosis grades as determined by pathology. Fig. 3B shows the DOPU value profiles for different fibrosis levels based on pathology from 6 donor livers, with an inset plot indicating that higher fibrosis levels correspond to higher DOPU values. A linear relationship was found between the average DOPU values and pathology scores, indicating that hepatic tissues with higher pathology scores have increased DOPU values (Fig. 3C).

**Fig. 3.**
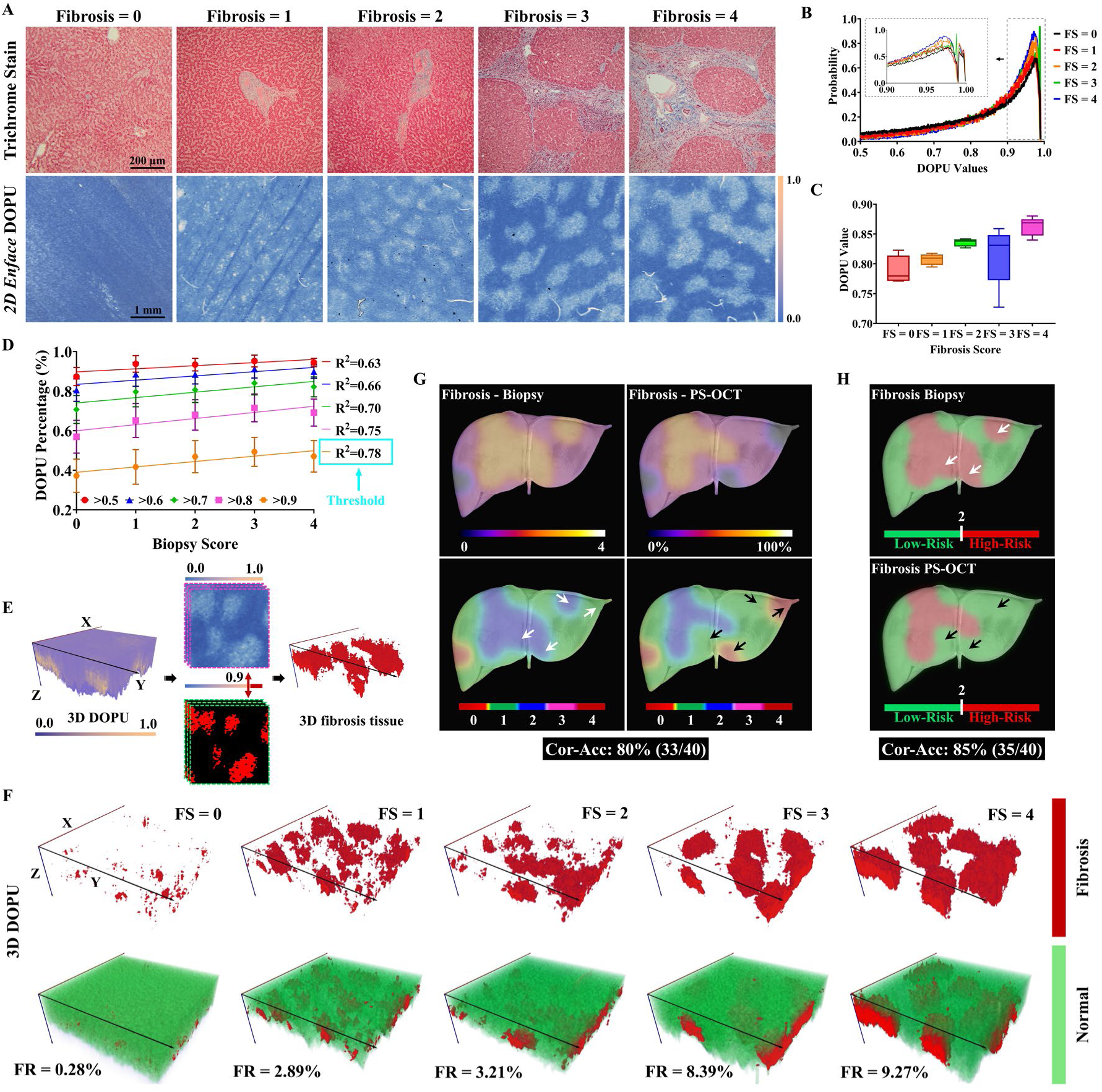
Recognition and quantification of hepatic fibrosis from PS-OCT DOPU images for viability evaluation. (A) Representative pathology and PS-OCT DOPU images at different fibrosis levels. (B) Profile of DOPU values at different fibrosis degrees. (C) Distribution characteristics of DOPU values and pathology scores. (D) Linear fitting between DOPU percentage and pathology scores for threshold screening. (E) 3D reconstruction of fibrosis from DOPU images. (F) Representative 3D fibrosis structures at different pathological fibrosis scores. (G) Correlation and comparison of fibrosis distribution over entire liver surfaces between DOPU and pathology. (H) Comparison of fibrosis regions at Low-Risk and High-Risk between DOPU and pathology under clinical and fitting-based thresholds. FS, fibrosis score. Cor-Acc, correlation accuracy. FR, fibrosis ratio.

To further correlate PS-OCT DOPU images with pathology scores, we performed a linear fitting to correlate DOPU percentages extracted using different thresholds with pathology scores. An optimal threshold of 0.9 was identified for fibrosis extraction from PS-OCT DOPU images, based on the highest R-square value of the linear fitting (Fig. 3D). Using this threshold, we extracted the 3D fibrosis structure from each PS-OCT DOPU dataset for quantification (Fig. 3E and Movie S2). Representative 3D fibrosis microstructures, corresponding to various pathology scores, and their calculated percentages within the 3D hepatic tissues are shown in Fig. 3F. These data indicate that tissues with higher fibrosis pathology scores exhibit more fibrosis as extracted from PS-OCT DOPU images.

In Fig. 3G, we present the distribution characteristics of fibrosis in a representative donor liver, mapped using both PS-OCT imaging and pathology. The top images depict the heterogeneous distribution of fibrosis as determined by DOPU and pathological quantification, while the bottom images show varying fibrosis levels based on DOPU and pathology scores. Our results revealed a strong correlation between PS-OCT DOPU imaging and pathology, with an 80% correlation accuracy across 33 out of 40 sites. However, discrepancies were noted in four specific regions of the liver surface, marked by white and black arrows. When applying current clinical standards to distinguish low-risk from high-risk regions, the correlation accuracy between PS-OCT and pathology were 85% (35 out of 40), as shown in Fig. 3H.

### Hepatic Inflammation Quantification and Classification by PS-OCT and Texture Feature

We extracted 2,484 texture features for each scanning site across the entire liver and compared these features at different pathologic inflammation scores using a volcano plot (Fig. 4A). Most features showed significant differences across different inflammatory liver tissues, although the degree of difference was relatively low. Following feature selection using a random forest model, we obtained the spatial distribution of the top three most effective features for classifying specific inflammation scores and regions (Fig. 4B). The total set of effective features and their corresponding weights are shown in Fig. S3. These three effective features demonstrated spatial distribution differences when classifying varying inflammation scores and regions. The classification of inflammation regions, as opposed to specific scores, showed a higher level of significance in spatial distribution.

**Fig. 4.**
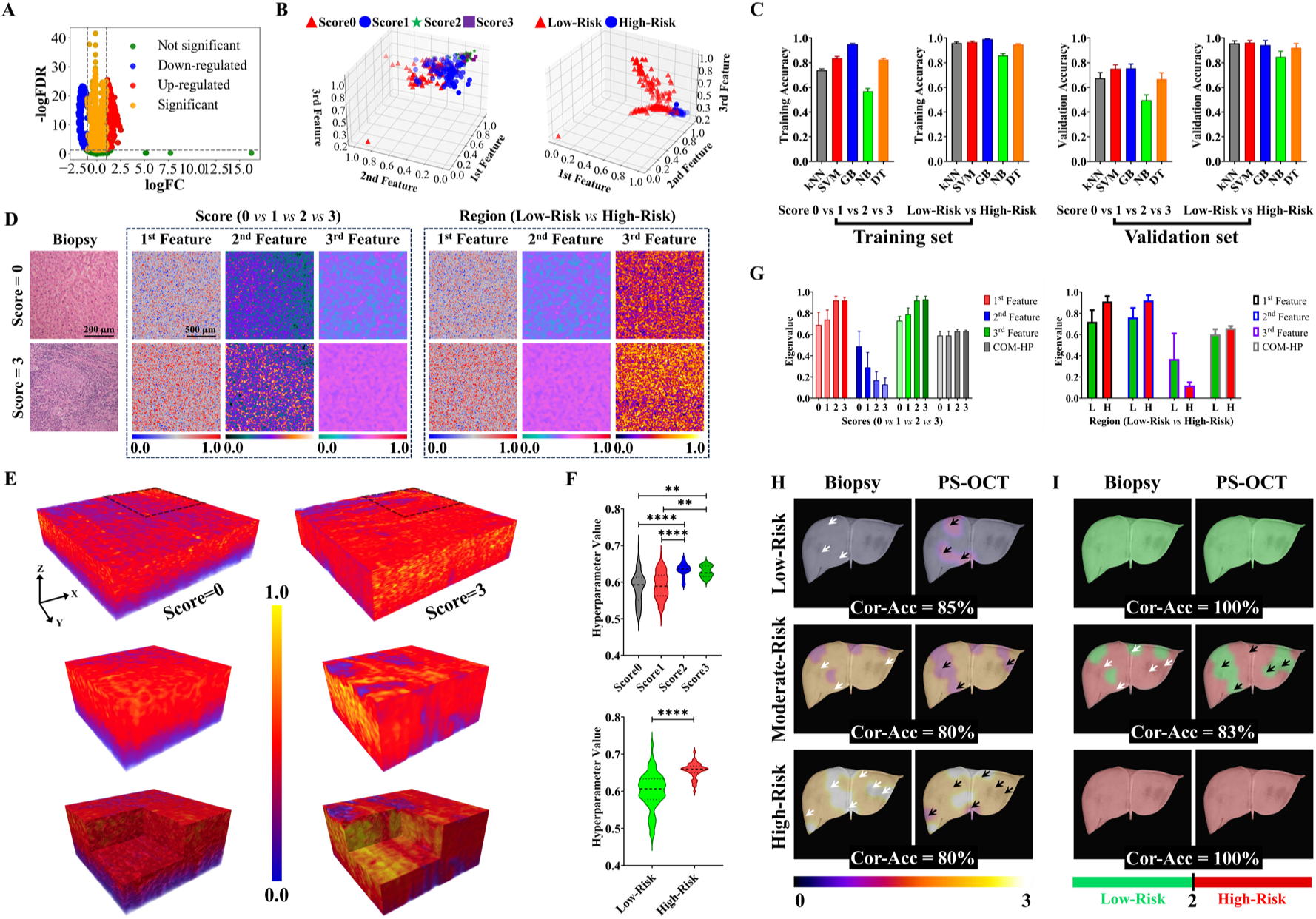
Extraction and quantification of hepatic inflammation by texture features from PS-OCT intensity images. (A) Texture feature distribution of hepatic inflammation classification. (B) The top three effective texture features of hepatic inflammation classification under the random forest model. (C) Classification accuracy of machine learning models in hepatic inflammation classification. (D) Representative of the top three feature images for hepatic inflammation classification and the corresponding histology images. (E) 3D reconstruction of the top three features and the composite hyperparameter with none inflammation (score=0) and severe inflammation (score=3). (F) The eigenvalue distribution of the composite hyperparameter for specific scores and regions of the hepatic inflammation. (G) Eigenvalue distributions of the top three features and the composite hyperparameters in classifying specific scores and regions of hepatic inflammation. (H) Correlation and comparison of inflammation distribution over entire liver surface between PS-OCT and biopsy. (I) Comparison of inflammation regions at Low-Risk and High-Risk between PS-OCT and pathology under clinical and machine learning classification thresholds. Cor-Acc, correlation accuracy. In the classification of inflammation scores (0, 1, 2, 3), 1^st^ Feature is WP_coif1_hha_mean, 2^nd^ Feature is FDTA_HurstCoeff_2, 3^rd^ Feature is WP_coif1_hdv_mean. In the classification of inflammation regions (Low-Risk and High-Risk), 1^st^ Feature is WP_coif1_hha_mean, 2^nd^ Feature is WP_coif1_hdv_mean, 3^rd^ Feature is Correlogram_31_5_hd.

Fig. 4C presents a comparison of classification accuracy across different machine learning models, where the SVM model achieved the highest accuracy for both inflammation scores and regions. Table 2 provides details on classification accuracy and ROC-AUC values. The ROC-AUC and distribution characteristics of the top two effective features for inflammation classification are shown in Fig. S4. In Fig. 4D, 2D PS-OCT texture feature images (specifically, the top three effective features) are compared with corresponding histology images at various inflammation scores, revealing notable differences in texture patterns and distributions. Table 3 lists the eigenvalue differences of the top three effective features in classifying specific inflammation scores.

**Table 2.** The comparison of classification accuracy for machine learning models in hepatic inflammation classifications. Tra, training set. Acc, accuracy. Str, standard error. Val, validation set. ROC, receiver-operating characteristic curve. AUC, area under the curve.

| Score 0 vs 1 vs 2 vs 3 |  |  |  |  |  |  |
| --- | --- | --- | --- | --- | --- | --- |
|  | Tra-Acc | Tra-Str | Val-Acc | Val-Str | ROC-AUC | Str |
| **kNN** | 0.739 | 0.011 | 0.675 | 0.045 | 0.825 | 0.064 |
| **SVM** | 0.836 | 0.015 | 0.751 | 0.032 | 0.867 | 0.035 |
| **GB** | 0.951 | 0.006 | 0.755 | 0.035 | 0.833 | 0.031 |
| **NB** | 0.569 | 0.022 | 0.496 | 0.043 | 0.756 | 0.049 |
| **DT** | 0.825 | 0.009 | 0.667 | 0.052 | 0.794 | 0.073 |
| Low-Risk vs High-Risk |  |  |  |  |  |  |
| **kNN** | 0.959 | 0.009 | 0.957 | 0.021 | 0.973 | 0.031 |
| **SVM** | 0.967 | 0.008 | 0.963 | 0.017 | 0.988 | 0.009 |
| **GB** | 0.991 | 0.004 | 0.943 | 0.036 | 0.984 | 0.015 |
| **NB** | 0.859 | 0.016 | 0.847 | 0.046 | 0.926 | 0.057 |
| **DT** | 0.948 | 0.006 | 0.922 | 0.035 | 0.882 | 0.063 |

**Table 3.** Texture feature eigenvalues and composite hyperparameter values of hepatic inflammation. COM-HP, composite hyperparameter.

|  | Score 0 vs 1 vs 2 vs 3 |  |  |  | Low-Risk vs High-Risk |  |
| --- | --- | --- | --- | --- | --- | --- |
|  | Score 0 | Score 1 | Score 2 | Score 3 | Low-Risk | High-Risk |
| **1<sup>st</sup> Feature** | 0.69±0.12 | 0.74±0.09 | 0.92±0.04 | 0.92±0.03 | 0.72±0.11 | 0.91±0.05 |
| **2<sup>nd</sup> Feature** | 0.49±0.14 | 0.29±0.14 | 0.17±0.08 | 0.13±0.06 | 0.76±0.09 | 0.92±0.05 |
| **3<sup>rd</sup> Feature** | 0.73±0.11 | 0.79±0.06 | 0.92±0.04 | 0.93±0.03 | 0.37±0.24 | 0.12±0.03 |
| **COM-HP** | 0.59±0.04 | 0.59±0.04 | 0.63±0.02 | 0.63±0.01 | 0.60±0.05 | 0.66±0.02 |

To visualize all effective texture features in inflammation classification in 3D, we reconstructed the 3D texture structure using composite hyperparameters, highlighting the spatial distribution of inflammation (Movie S3). Fig. 4E illustrates that liver tissues without inflammation (score = 0) and those with severe inflammation (score = 3) exhibit substantial differences in spatial texture patterns and distributions, which correspond to histology findings in Fig. 4D. Fig. 4F compares composite hyperparameters among different inflammation scores (0, 1, 2, 3) and between low-risk and high-risk regions. Significant differences were observed in composite hyperparameter values when comparing scores 0 vs. 2 (P < 0.0001), 0 vs. 3 (P < 0.01), 1 vs. 2 (P < 0.0001), and 1 vs. 3 (P < 0.01). Additionally, there was a significant difference between the composite hyperparameters of low-risk and high-risk regions (P < 0.0001).

Fig. 4G and Table 3 show substantial differences and trends in the eigenvalues of the top three effective features and composite hyperparameters. Specifically, inflammation scores 0 and 1 are significantly different from scores 2 and 3. Moreover, the low-risk and high-risk regions exhibit significant differences based on the top three effective features and composite hyperparameters. In Fig. 4H, we display the inflammation score distribution of three representative donor livers, comparing PS-OCT-imaging inflammation scores with corresponding biopsy scores. These livers are categorized as low-risk (scores 0, 1), moderate-risk (scores 1, 2), and high-risk (scores 2, 3) based on the biopsy inflammation scores over the entire liver. A strong correlation was observed between PS-OCT and biopsy results for hepatic inflammation score evaluation. Fig. 4I compares the clinical standard (pathology) with the SVM-based texture feature standard in differentiating hepatic inflammation regions, where PS-OCT showed a stronger correlation with biopsy findings.

### Hepatic Necrosis Quantification and Classification by PS-OCT and Texture Feature

We also extracted 2,484 texture features to compare and classify hepatic necrosis across specific biopsy scores and regions. As shown in Fig. 5A, most extracted texture features displayed significant differences in hepatic necrosis with specific scores, although the degree of difference was relatively low. The total effective features and their corresponding weights are presented in Fig. S5. The top three most effective features, selected using a random forest model, were used to visualize the spatial distribution for classifying hepatic necrosis by specific scores and regions. Classification of necrotic regions showed greater significance than classification by necrosis scores (Fig. 5B).

**Fig. 5.**
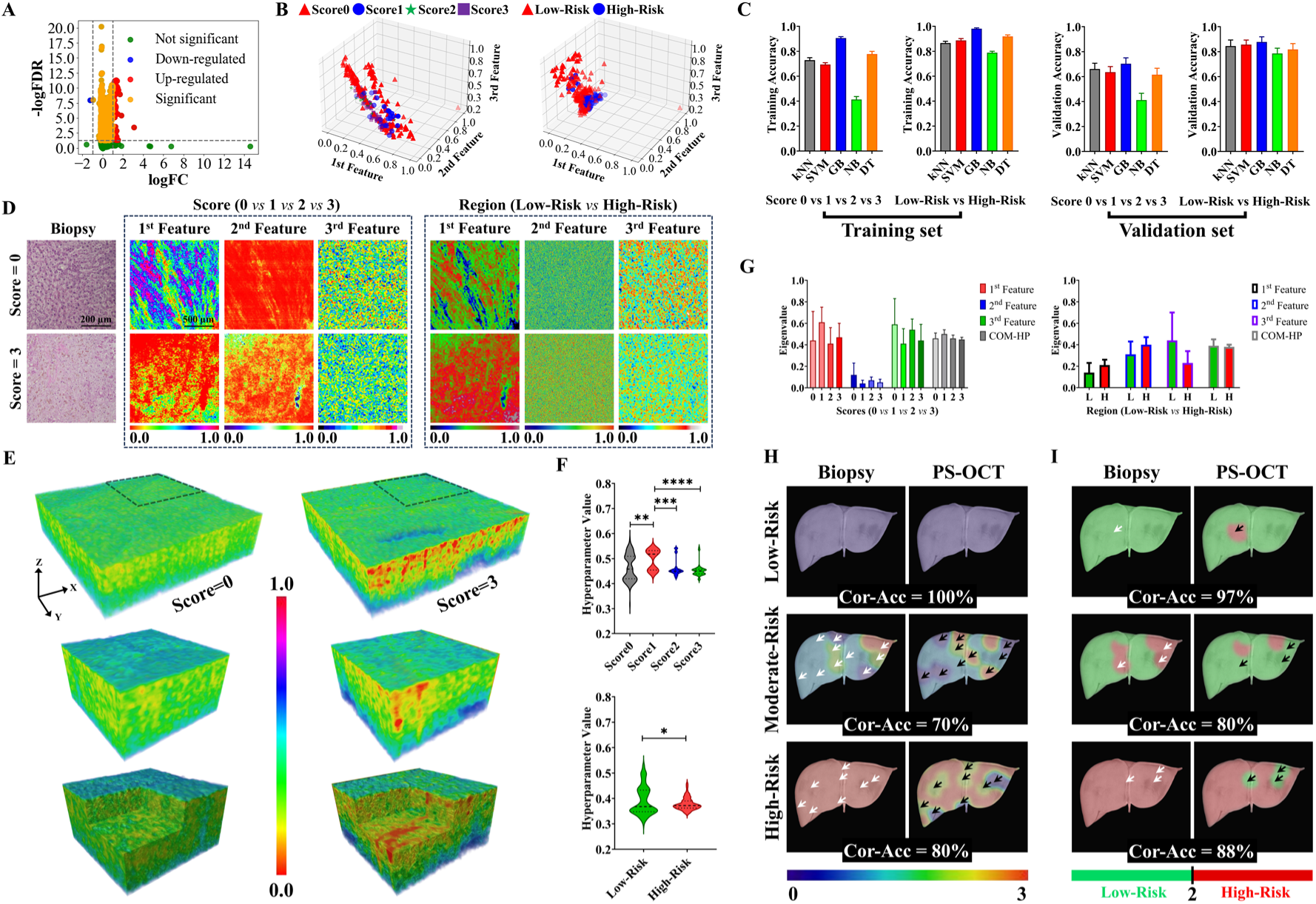
Extraction and quantification of hepatic necrosis by texture features from PS-OCT intensity images. (A) Texture feature distribution of hepatic necrosis classification. (B) The top three effective texture features of hepatic necrosis classification under the random forest model. (C) Classification accuracy of machine learning models in hepatic necrosis classification. (D) Representative of the top three feature images for hepatic necrosis classification and the corresponding histology images. (E) 3D reconstruction of the top three features and the composite hyperparameter with no necrosis (score=0) and severe necrosis (score=3). (F) The eigenvalue distribution of the composite hyperparameter for specific scores and regions of the hepatic necrosis. (G) Eigenvalue distributions of the top three features and the composite hyperparameters in classifying specific scores and regions of hepatic necrosis. (H) Correlation and comparison of necrosis distribution over entire liver surface between PS-OCT and biopsy. (I) Comparison of necrosis regions at Low-Risk and High-Risk between PS-OCT and pathology under clinical and machine learning classification thresholds. Cor-ACC, correlation accuracy. In the classification of necrosis scores (0, 1, 2, 3), 1^st^ Feature is Smr1, 2^nd^ Feature is GLRLM_Short Low_Gray_Level_Emphasis, 3^rd^ Feature is Correlogram_14_5_ht. In the classification of necrosis regions (Low-Risk and High-Risk), 1^st^ Feature is WP_coif1_aav_std, 2^nd^ Feature is LBP_R_3_P_24_entropy, 3^rd^ Feature is Correlogram_0_8_ht.

Fig. 5C and Table 4 demonstrate that the Gradient Boosting (GB) model achieved the highest classification accuracy for hepatic necrosis, both at the level of specific scores and regions. The corresponding ROC-AUC and the distribution characteristics based on the top two effective features are detailed in Fig. S6. To assess the classification performance of hepatic necrosis from PS-OCT images, we compared images of the top effective feature with corresponding biopsy images (Fig. 5D). The texture feature images showed varying texture patterns and distributions according to different necrosis scores, with the eigenvalues provided in Table 5.

**Table 4.** The comparison of classification accuracy for machine learning models in hepatic necrosis classifications. Tra, training set. Acc, accuracy. Str, standard error. Val, validation set. ROC, receiver-operating characteristic curve. AUC, area under the curve.

| Score 0 vs 1 vs 2 vs 3 |  |  |  |  |  |  |
| --- | --- | --- | --- | --- | --- | --- |
|  | Tra-Acc | Tra-Str | Val-Acc | Val-Str | ROC-AUC | Str |
| **kNN** | 0.728 | 0.020 | 0.660 | 0.047 | 0.777 | 0.039 |
| **SVM** | 0.694 | 0.014 | 0.635 | 0.045 | 0.812 | 0.053 |
| **GB** | 0.906 | 0.011 | 0.704 | 0.047 | 0.840 | 0.045 |
| **NB** | 0.413 | 0.024 | 0.412 | 0.055 | 0.740 | 0.059 |
| **DT** | 0.778 | 0.022 | 0.616 | 0.052 | 0.686 | 0.073 |
| Low-Risk vs High-Risk |  |  |  |  |  |  |
| **kNN** | 0.866 | 0.014 | 0.845 | 0.048 | 0.871 | 0.039 |
| **SVM** | 0.888 | 0.014 | 0.857 | 0.036 | 0.913 | 0.037 |
| **GB** | 0.981 | 0.007 | 0.878 | 0.041 | 0.940 | 0.033 |
| **NB** | 0.789 | 0.012 | 0.786 | 0.042 | 0.890 | 0.042 |
| **DT** | 0.919 | 0.012 | 0.818 | 0.047 | 0.814 | 0.061 |

**Table 5.** Texture feature eigenvalues and composite hyperparameter values of hepatic necrosis. COM-HP, composite hyperparameter.

|  | Score 0 vs 1 vs 2 vs 3 |  |  |  | Low-Risk vs High-Risk |  |
| --- | --- | --- | --- | --- | --- | --- |
|  | Score 0 | Score 1 | Score 2 | Score 3 | Low-Risk | High-Risk |
| **1<sup>st</sup> Feature** | 0.44±0.27 | 0.61±0.14 | 0.41±0.15 | 0.47±0.13 | 0.14±0.09 | 0.21±0.05 |
| **2<sup>nd</sup> Feature** | 0.12±0.11 | 0.04±0.03 | 0.07±0.03 | 0.05±0.03 | 0.31±0.12 | 0.40±0.07 |
| **3<sup>rd</sup> Feature** | 0.59±0.24 | 0.41±0.14 | 0.54±0.10 | 0.44±0.15 | 0.44±0.26 | 0.23±0.11 |
| **COM-HP** | 0.46±0.05 | 0.50±0.04 | 0.46±0.03 | 0.45±0.02 | 0.39±0.06 | 0.38±0.02 |

Fig. 5E and Movie S4 present the 3D reconstruction of hepatic necrosis based on composite hyperparameters, highlighting the differences between hepatic tissues without necrosis (score = 0) and those with severe necrosis (score = 3). We observed substantial differences in spatial texture patterns and distributions when examining the top three effective features and composite hyperparameters. In Fig. 5F, the statistical characteristics of composite hyperparameter values are shown for specific necrosis scores and regions. Significant differences were found among the comparisons of scores 0 vs. 1 (P < 0.01), 1 vs. 2 (P < 0.001), and 1 vs. 3 (P < 0.0001). Additionally, the composite hyperparameter distribution differed significantly between low-risk and high-risk regions.

To further explore texture differences and trends across necrosis scores and regions, we compared the eigenvalues of the top three effective features and composite hyperparameters. Our results indicate that hepatic necrosis in low-risk and high-risk regions shows substantial differences in texture features (Fig. 5G and Table 5). However, although significant differences were noted for specific necrosis scores, no clear trend emerged. We mapped the distribution of specific hepatic necrosis scores across three representative donor livers, comparing the correlations between PS-OCT findings and corresponding biopsy results (Fig. 5H). These livers were categorized as low-risk (scores 0, 1), moderate-risk (scores 1, 2), and high-risk (scores 2, 3) based on pathological necrosis scores throughout the liver. A strong correlation between PS-OCT and biopsy results was observed for hepatic necrosis score evaluation in these livers. Finally, in Fig. 5I, we compared the current clinical standard (biopsy) and the GB-based texture feature standard (PS-OCT) in distinguishing hepatic necrosis regions. We found that PS-OCT and biopsy demonstrated a stronger correlation in the evaluation of hepatic necrosis regions than in the evaluation of specific necrosis scores.

### Comprehensive Evaluation of Donor Liver Viability by PS-OCT Imaging

We conducted a comprehensive viability evaluation of representative donor livers using PS-OCT imaging. Fig. 6A displays five donor livers with suggested decisions based on these evaluations. A radar chart illustrates the evaluation performance of each liver, considering steatosis, fibrosis, inflammation, and necrosis from the inner to the outer rings. Based on specific parameter scores and the identified regions across each liver, we proposed suggestive decisions. The absolute high-risk parameter (AHRP), defined as any high-risk hepatic parameter (score ≥ 2) covering all scanned sites across the entire liver, was used to assess overall liver viability.

**Fig. 6.**
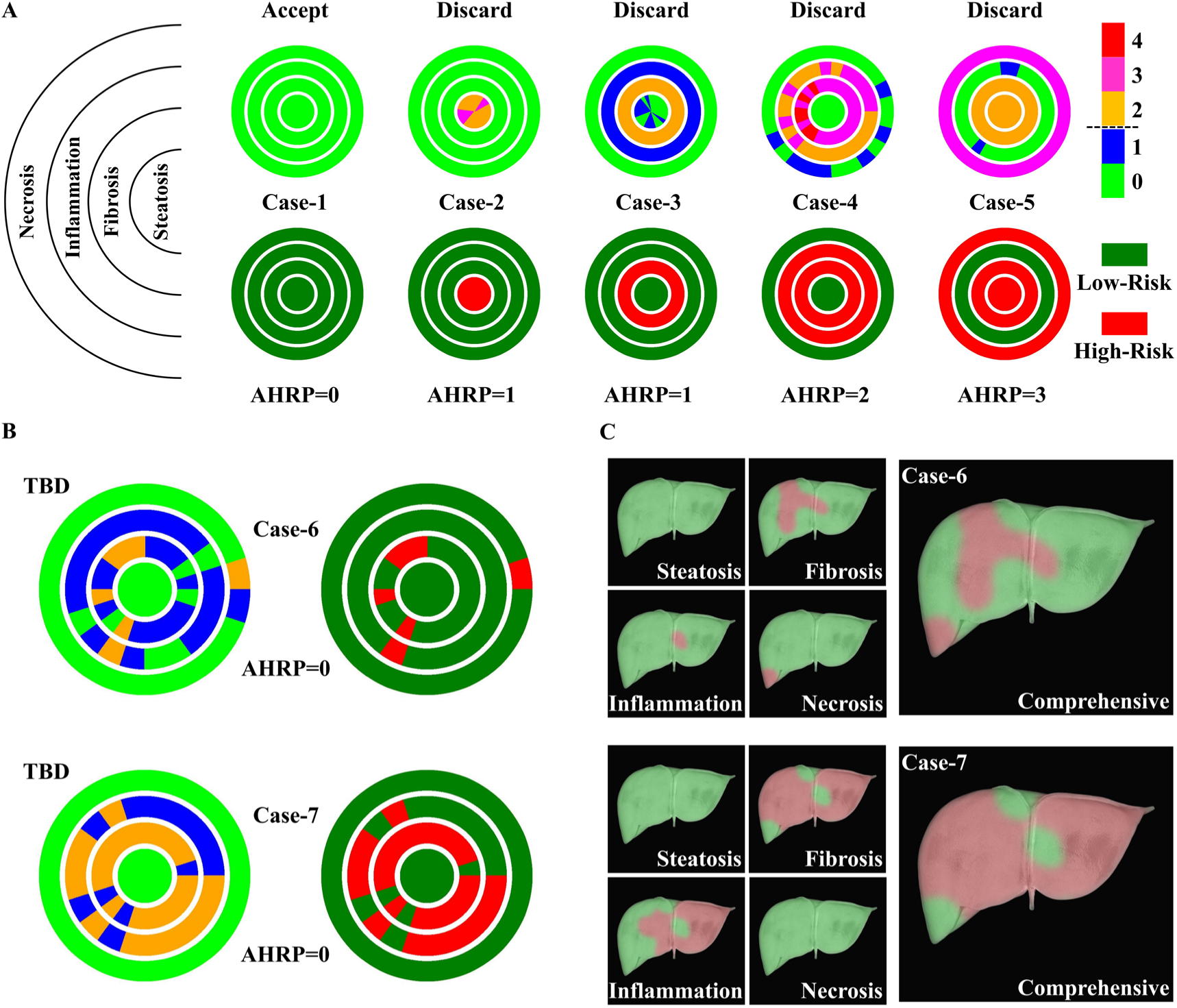
The comprehensive assessment of donor liver viability with hepatic steatosis, fibrosis, inflammation, and necrosis by PS-OCT imaging. (A) Suggested decisions for multiple donor liver cases under different conditions. (B) Representative donor liver cases showing significant viability heterogeneity, highlighting the request for a final decision. (C) Distribution of hepatic heterogeneity under various comprehensive and single-parameter evaluations. AHRP, absolute high-risk parameter. TBD, to be decided.

The liver in Case-1 was evaluated as acceptable for transplantation due to the absence of steatosis, fibrosis, inflammation, and necrosis. In contrast, Case-2 and Case-3 were recommended for discard due to the presence of steatosis and fibrosis throughout the entire liver, respectively. Case-4 exhibited fibrosis and inflammation over the entire liver, leading to a suggested discard. Case-5 showed severe conditions, with steatosis, fibrosis, and necrosis present across the entire liver, resulting in a recommendation for discard. In these cases, PS-OCT imaging results aligned with biopsy decisions for four cases (Cases-2, -3, - 4, and -5).

Interestingly, we identified two cases with partial high-risk evaluations of specific parameters. As shown in Fig. 6B, Case-6 exhibited localized fibrosis, inflammation, and necrosis, whereas Case-7 demonstrated fibrosis and inflammation across most regions, though not throughout the entire liver. The distribution of each hepatic parameter across the whole liver was mapped for both cases, and a comprehensive evaluation map was created based on all four parameters to assess liver viability. We found that the majority of regions in Case-6 were deemed low-risk, while most regions in Case-7 were identified as requiring discard (Fig. 6C). Given the absence of AHRP in both cases, we suggested that final decisions be made by clinicians based on the comprehensive evaluations.

## DISCUSSION

The shortage of available donor livers, driven by the high demand for liver transplantation and persistently high organ discard rates, underscores the urgent need for a reliable pre-transplantation liver viability evaluation tool to help increase transplant numbers, optimize transplant outcomes, and decrease discard rates. We propose using PS-OCT as an imaging modality to perform a comprehensive and nearly real-time viability evaluation by scanning the entire donor liver. To validate the PS-OCT images, we correlated the imaging results with pathological analysis, which is the current gold standard for assessing pretransplant donor liver viability. Our pathologic evaluation of the entire donor liver revealed significant regional heterogeneity in degrees of steatosis, fibrosis, inflammation, and necrosis across the liver surface (Fig. 1). These heterogeneities arise partly because hepatocytes in different liver zones vary in enzyme expression and subcellular structures, leading to functional and metabolic differences across the liver at the tissue level (*47, 48*). Additionally, hepatic ischemia-reperfusion injury is related to this intrinsic heterogeneity, as certain zones may be more vulnerable to injury (*49*). Consequently, biopsy from multiple sites is often necessary to accurately assess hepatic parameters, predict functional potential, and evaluate viability (*50–52*). Our histopathological examination of the donor liver surface revealed these patterns of heterogeneity, aligning with previous reports and reinforcing the importance of multi-site pathological analysis for a comprehensive viability assessment.

Conventional pathology, which relies on analyzing one or two tissue samples, may introduce significant sampling error and negatively impact the accuracy of viability evaluations for the entire donor liver. Furthermore, rapid liver assessment via pathology is not always feasible in hospitals due to the time required for processing, and doing enough biopsies to avoid sampling error is unlikely to be feasible or advisable. The reliability of pathologic findings at different parameter cutoffs or between various pathologists is also a factor that cannot be ignored, as significant inter-observer variability is common (*53–55*). Additionally, liver biopsy are invasive and associated with risks, such as bleeding, despite the small sample size (*56*). Our findings align with current clinical concerns regarding the limitations of graft outcome prediction by the LDRI and the gradings in liver viability assessments based on pathology (*13, 14, 57*). In PS-OCT images, through noninvasive and noncontact scanning across the entire donor liver, we effectively address these clinical problems by providing comprehensive qualification and quantification of steatosis, fibrosis, inflammation, and necrosis. These four parameters are key indicators for evaluating liver viability for transplantation in pathology assessment (*16, 24–27*). The strong correlation between PS-OCT and pathologic evaluations suggests that PS-OCT, in conjunction with routine pathologic evaluation, can accurately assess donor liver viability and has the potential to achieve fully automated imaging evaluations in the future. Functional tests utilizing biomarkers produced during normothermic machine perfusion is another effective liver viability assessment, however, less than 10% of liver grafts currently undergo machine perfusion due to its complexity and high cost (*58*). Consequently, the majority (∼90%) of liver viability and graft assessments still rely on pathology, highlighting the importance of PS-OCT imaging for liver assessment. Moreover, the PS-OCT imaging system is compatible with existing *ex vivo* liver perfusion devices. This allows the microstructural information provided by PS-OCT imaging to be combined with the functional tests collected during ex-vivo perfusion. Our PS-OCT system is based on the widely used spectral-domain OCT, which has proven to be an operationally practical tool for kidney imaging in ongoing clinical trials (*59*). PS-OCT could easily be adopted and utilized by clinicians for liver transplantation trials in the future.

Hepatic steatosis is the most critical parameter in liver viability evaluation, with size and density serving as key indicators for grading and quantification (*21, 57, 60*). Histologically, steatosis is categorized as microvesicular or macrovesicular. However, current viability evaluations often focus primarily on macrovesicular steatosis (*21*). The combined percentage of macro- and micro-steatosis is typically used to assess liver transplant feasibility (*61–63*), with degrees of steatosis classified as mild (<30%), moderate (30%-60%), or severe (>60%) (*15, 64, 65*). A threshold of 30%-33% macrovesicular steatosis generally determines suitability for use in transplantation, with higher levels indicating unsuitability (*21, 61, 66, 67*). However, current pathology grading only provides a quantitative estimation of two-dimensional (2D) steatosis, limited by the constraints of histological staining of liver tissues. In addition to the limitation of regional evaluation from single-sample histology staining, the specific methods used for quantitative evaluation of pathology stains can also introduce errors in liver tissue grading (*21*). Fortunately, our results revealed that PS-OCT can directly detect steatosis due to the fat droplet microstructure can be effectively identified by OCT imaging (*68*). PS-OCT images enable direct identification and segmentation of steatosis for quantification, as shown in Fig. 2. More importantly, PS-OCT allows for 3D segmentation of steatosis, providing a more reliable measurement of spatial size and volume density compared to traditional pathology. Since PS-OCT quantifies steatosis in 3D, there are quantitative and qualitative differences in steatosis density measurements between PS-OCT and traditional pathology. This discrepancy necessitates the development of a dedicated standard for PS-OCT to quantitatively evaluate hepatic steatosis. To establish this dedicated standard correlation with the clinical threshold of 33% (*21*), we used distribution statistics, a widely applied method (*69*), to generate a threshold for evaluating hepatic steatosis. With the established steatosis density threshold (0.04%), PS-OCT images not only reveal the heterogeneous distribution of steatosis across the entire donor liver but also display a strong correlation (85%) with the clinical pathology evaluation.

Hepatic fibrosis is a parameter of liver viability evaluation and is associated with hepatic inflammation and necrosis (*24–26*). Hepatic fibrosis is quantitatively evaluated by portal tract fibrosis which refers to the development of fibrous (scar) tissue within the portal tracts of the liver (*70–72*). Different stages of fibrosis can be identified by the progression and extent of fibrous tissue in the liver (*73, 74*). Since fibrosis produces birefringence by altering the polarization state of light (*75*), PS-OCT polarization images can detect these changes to measure the degree of fibrosis (Fig. 3 and Fig. S2). Our results show that the different stages of fibrosis identified in pathology images can also be recognized in corresponding PS-OCT DOPU images. The correlation between different fibrosis stages in PS-OCT and pathology follows a linear relationship, enabling the development of a grading system for hepatic fibrosis detected on PS-OCT DOPU images. Using this grading system, our PS-OCT achieved an 80% correlation accuracy with pathology for specific fibrosis scores and an 85% correlation accuracy for liver viability evaluation. This precise one-to-one alignment has not been achieved with other clinical imaging modalities, including ultrasound, computed tomography (CT), or magnetic resonance imaging (MRI) (*76–78*). Although fluorescence imaging and two-photon microscopy can detect different stages of hepatic fibrosis, their clinical utility for liver viability evaluation is limited by the need for imaging dyes, low tissue penetration, and complex sample processing requirements (*79–81*). The noninvasive, high-resolution capabilities of PS-OCT make it an ideal tool for evaluating hepatic fibrosis across the entire liver.

Hepatic inflammation and necrosis are two key tissue alterations resulting from the injury and death of hepatocytes. However, these changes do not cause significant microstructural alterations. As a result, PS-OCT images cannot directly quantify tissue changes associated with inflammation and necrosis. Although these tissue alterations cannot be detected directly, they alter the scattering properties of light, allowing changes in pixel intensity to be measured and used to monitor these alterations (*82*). Texture features have been widely used to extract pixel pattern information for remote sensing and medical imaging to segment and classify specific regions of the donor liver (*83–86*). To monitor these pixel intensity changes, we use texture features to classify pixel patterns for quantifying hepatic inflammation and necrosis. By applying machine learning models, we identified effective features from 27 texture parameters to classify specific scores and risk categories for hepatic inflammation and necrosis. Since 3D PS-OCT intensity images cannot directly distinguish liver tissue at different stages of hepatic inflammation and necrosis, we reconstructed 3D hyperparameter structures by convoluting-summing all effective features. This approach effectively visualizes liver tissue with and without inflammation or necrosis. The feature eigenvalues reveal significant differences across various stages of inflammation and necrosis which cannot be directly quantified by PS-OCT images (Fig. 4 and 5). When validated by pathologic results, PS-OCT shows a strong correlation in classifying hepatic inflammation and necrosis. Notably, PS-OCT demonstrates a relatively stronger correlation with pathologic data when classifying risk categories (low-risk and high-risk) for hepatic inflammation and necrosis than when classifying specific scores. Therefore, it indicates that PS-OCT images combining texture features have a higher sensitivity to classify the risk categorie across the entire liver. We noticed that the top three effective features and the hyperparameter in hepatic necrosis classifications have a fluctuation alteration of feature eigenvalues in evaluation scores. We speculate that advanced stages of hepatic necrosis (scores 3 and 4), which indicate submassive and massive hepatocyte death, may produce structural features indicative of early stages of necrosis (scores 0 and 1) (*87*). This overlap could potentially disrupt the expected trend changes in texture features.

PS-OCT shows a strong correlation with pathology in classifying evaluation scores and risk categories, particularly achieving high accuracy in predicting final decisions for donor livers with absolute high-risk parameters (AHRP). This finding suggests that PS-OCT may have the potential to replace traditional pathologic analysis for a rapid and comprehensive pretransplant viability evaluation of the entire deceased donor liver. The possible utility of PS-OCT in decreasing the discard rate of higher risk donor livers and to expand the donor pool has not been confirmed. Fortunately, we provide evidence that PS-OCT can also provide the global distribution characteristics of risk categories across the entire liver. For those marginal livers without AHRP, PS-OCT allows the visualization of risk categories (low-risk *vs* high-risk) to offer a more comprehensive reference to help inform clinical decision-making. Some livers with high-risk risk factors may still be suitable for transplantation if the risks are limited and if they are adequately assessed (*88*). Our results show that donor livers with limited high-risk regions can be further assessed by clinicians to make final decisions for or against use (Fig. 6B), potentially reducing the discard rate of marginal livers and expanding the donor pool. Additionally, with PS-OCT’s ability to evaluate viability across the entire liver, marginal livers with limited high-risk factors may still be suitable for partial liver transplantation (*89, 90*). It has the potential to help patients in urgent need of transplantation or those who have been on the waiting list for an extended period. Still, reports have documented cases of graft failure following transplantation due to the use of high-risk marginal livers (*22, 91*). Therefore, comprehensive viability evaluation of the entire liver with PS-OCT can help avoid the use of marginal livers with extensive high-risk regions, potentially reducing the risk of primary graft nonfunction (PNF) or early allograft dysfunction (EAD).

Notably, we used 20 deceased donor livers to investigate the feasibility of PS-OCT for liver viability evaluation by quantifying hepatic steatosis, fibrosis, inflammation, and necrosis parameters. The correlation between pathology and PS-OCT for these parameters is based solely on our current sample size. Therefore, the quantitative evaluation and statistical analysis of hepatic steatosis, fibrosis, inflammation, and necrosis are limited to these 20 livers. As the sample size increases, the quantitative indexes for these parameters may fluctuate. It is also important to note that PS-OCT currently provides an imaging depth of approximately 1.0∼2.0 mm in liver tissues due to tissue scattering and laser source limitations. Nonetheless, our study represents a pioneering effort in applying OCT imaging to overcome the limitations of traditional histology, enabling a more comprehensive and reliable evaluation of liver tissues. A comparison of histopathologic scores between liver tissues at depths of 2 mm and ∼2 cm, conducted by three board-certified pathologists, revealed no significant differences in conventional pathology staining (Fig. S7). This finding demonstrates that the imaging depth achieved by PS-OCT in this study is sufficient to obtain effective depth information for liver tissue assessment. With advancements in OCT modalities, such as longer-wavelength laser sources (*92, 93*) or matrix imaging modes (*94*), PS-OCT has the potential to achieve much higher penetration depth, making it a promising tool for clinical liver assessment. On the other hand, the detection and quantification of hepatic fibrosis based on DOPU are based on the cumulative values due to the single input polarization state in our PS-OCT system (*95, 96*). As a result, the local polarization information cannot be quantitatively provided. However, since the local birefringence mode is directly proportional to the actual birefringent signal intensity per pixel, the cumulative polarization signal offers higher sensitivity compared to the local polarization signal (*97–99*).

A recent report demonstrated that a polarization state tracing method, which follows the polarization evolution in depth along the Poincare sphere, can extract the local birefringence signal using a single input polarization state (*100*). By applying differential calculations to account for the transmission differences between adjacent layers, our PS-OCT imaging has the potential to reveal local fibrosis information effectively in the future study. Moreover, it is important to note that no test is perfectly reliable in assessment of graft viability. While our PS-OCT imaging significantly enhances liver viability evaluation compared to traditional pathology, pathology staining may still be required in specific high-risk zones (as in Case-6, Fig. 6B) to support a final decision. As the imaging time for the entire liver is currently constrained by the field-of-view (FOV) of the PS-OCT system, we are exploring the use of a space-division multiplexing OCT system to simultaneously acquire several parallel imaging zones (*101, 102*), enabling wide-field imaging of liver tissues and significantly reducing imaging time. Because PS-OCT intensity and polarization imaging provide only structural information, we have not yet obtained functional viability assessments, such as lactate clearance analysis (*103*). However, advancements in dynamic OCT have enabled functional imaging of metabolism and inflammation in mouse livers (*104*), presenting a promising pathway for developing a dynamic PS-OCT system to evaluate the functional viability (e.g., metabolism) of *ex vivo* human livers in future studies. Furthermore, with the advent of machine perfusion to enhance organ viability assessment, optimize organ quality, and extend preservation time, Doppler OCT may hold promise in the assessment of the functional characteristics of blood flow within the liver (*105, 106*).

In this study, we explored the feasibility of using PS-OCT for comprehensive viability evaluation of pre-transplantation deceased donor livers. Through pathological analysis, we identified significant heterogeneity in hepatic steatosis, fibrosis, inflammation, and necrosis across the entire donor liver surface, underscoring the necessity for global viability evaluation. We demonstrated that PS-OCT images provide spatially quantitative information on these parameters throughout the liver. Utilizing texture features and machine learning models, PS-OCT enabled a more comprehensive viability assessment and visualization of its distribution. Our findings suggest that PS-OCT has the potential to reduce discard rates for high-risk livers, thereby expanding the donor pool. This study offers new insights into the application of PS-OCT imaging for improving the viability evaluation process in human liver transplantation.

## METHODS AND MATERIALS

### Study Design

This study examines the feasibility of PS-OCT for the pre-transplant assessment of deceased donor livers, as illustrated in Fig. 7. The procedural overview, delineated in Fig. 7A, encompasses the PS-OCT scanning process and the comprehensive hepatic biopsy examination conducted across the entire liver. Deceased donor livers were obtained from *LifeShare of Oklahoma*, a non-profit Organ Procurement Organization (OPO) exclusively dedicated to organ and tissue retrieval for transplantation purposes. Manual labeling with medical ink at 20-40 locations covered the entirety of the liver surface, as illustrated in Fig. 7B. Subsequently, a customized PS-OCT system scanned each labeled position to acquire intensity and polarization images, facilitating the quantification of microstructures and tissue properties, as demonstrated in Fig. 7C. After imaging, hepatic tissue sections from each labeled position underwent histological staining, including H&E, PAS, Trichrome, PASD, and Iron stains, for the viability assessment of hepatic tissues (Fig. 7D). Employing established histopathologic criteria, three board-certified pathologists utilized a scoring sheet based on prevailing clinical evaluation standards (Banff Criteria) to grade the donor liver histopathological features. (Fig. 7E). To assess the correlation between PS-OCT and biopsy results, total steatosis was extracted from PS-OCT intensity images to align with histological data (Fig. 7F). Fibrosis quantification from PS-OCT Degree of Polarization Uniformity (DOPU) images was correlated with biopsy findings (Fig. 7G). Additionally, texture features from PS-OCT intensity images were extracted for integration into machine learning models, aligning with the evaluation of inflammation (Fig. 7H) and necrosis (Fig. 7I) performance from histological stains. Through the mapping of hepatic steatosis, fibrosis, inflammation, and necrosis from both biopsy and PS-OCT, we quantified the correlation between these modalities by assessing matching accuracy across the entire liver.

**Fig. 7.**
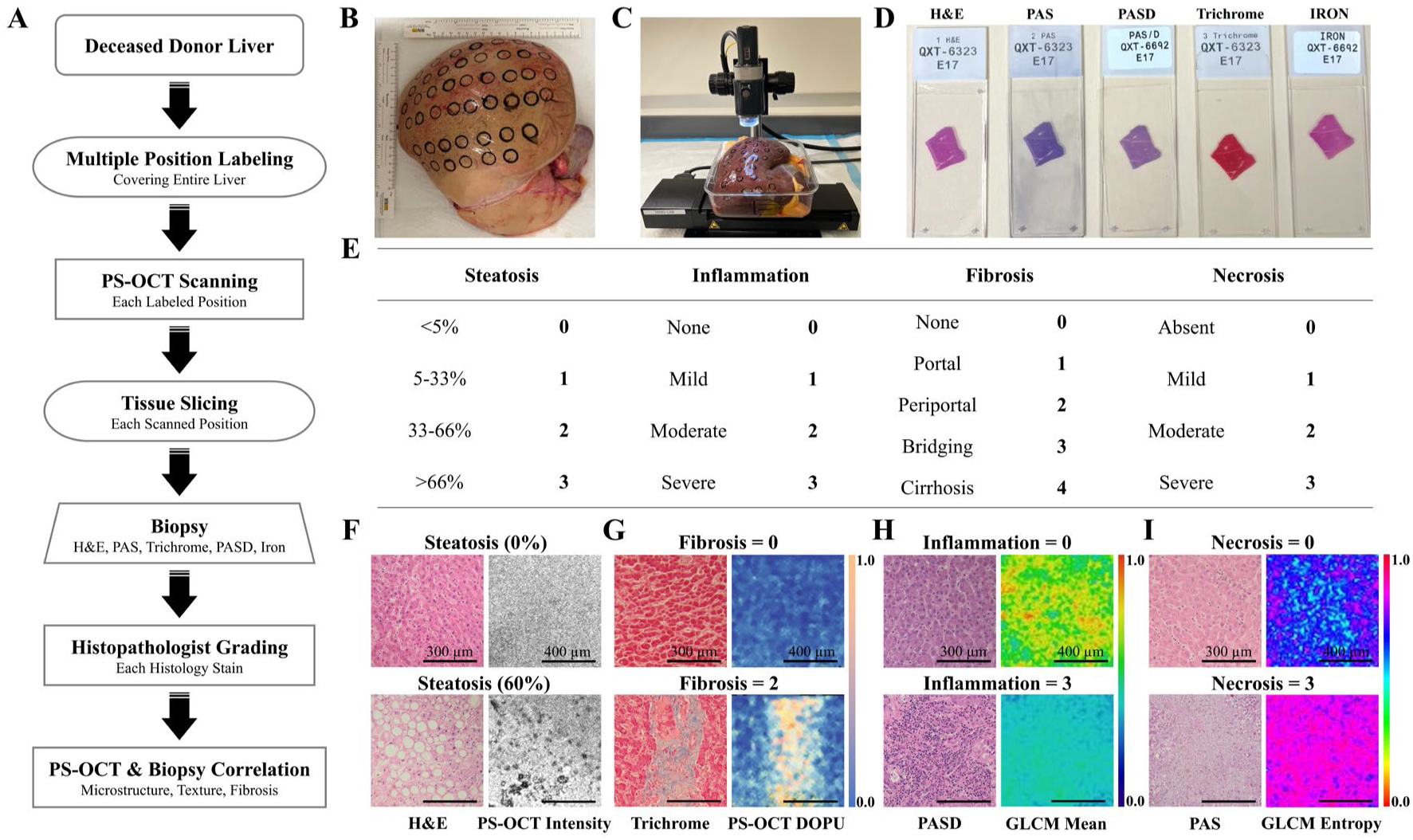
The framework of hepatic heterogeneity characterization and imaging parameters correlation between histopathology and PS-OCT based on deceased donor livers. (A) The flow chart of PS-OCT scanning and hepatic biopsy of the entire donor liver. (B) A representative deceased donor liver labeled with 40 imaging sites (dark circles). (C) The schematic of PS-OCT liver scanning. (D) Representative biopsies of hepatic tissues from a labeled region. (E) The score sheet of grading liver biopsies. (F) Representative H&E stains and PS-OCT intensity images of different degrees of steatosis. (G) Representative Trichrome stains and DOPU images of different degrees of fibrosis. (H) Representative PASD stains and PS-OCT texture images of different degrees of inflammation. (I) Representative PAS stains and PS-OCT texture images of different degrees of necrosis.

### Statistical Analysis

We used the Student’s *t*-test for comparing differences. A paired *t*-test was conducted to evaluate the correlation between biopsy results and PS-OCT measurements. An unpaired *t*-test was used to compare effective features across different scores and risk categories derived from PS-OCT texture feature extraction. A *P*-value of < 0.05 indicates a significant difference, while a *P*-value of ≥ 0.05 indicates that the difference is not significant. The correlation rates are calculated based on the true low-risk (TL), true high-risk (TH), false low-risk (FL), and false high-risk (FH). Specifically, the correlation accuracy, sensitivity, and specificity are obtained by:

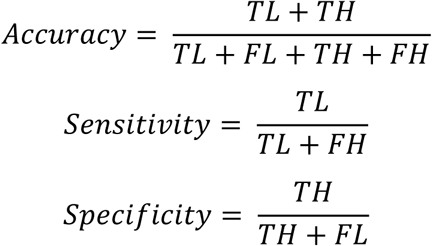

## Data Availability

All data produced in the present work are contained in the manuscript

## Acknowledgement

This work was supported by grants from the University of Oklahoma Health Sciences Center (3P30CA225520), National Science Foundation (OIA-2132161, 2238648, 2331409), National Institute of Health (R01DK133717), Oklahoma Center for the Advancement of Science and Technology (HR23-071), the medical imaging COBRE (P20 GM135009), and the Midwest Biomedical Accelerator Consortium (MBArC), an NIH Research Evaluation and Commercialization Hub (REACH). Histology service provided by the Tissue Pathology Shared Resource was supported in part by the National Institute of General Medical Sciences COBRE Grant P20GM103639 and National Cancer Institute Grant P30CA225520 of the National Institutes of Health. Financial support was provided by the OU Libraries’ Open Access Fund.

## Author Contributions

Conceptualization: QGT, YC, FY

Methodology: QGT, YC, FY, SP, PNM, NRB

Investigation: FY, QHZ, BMM, CW, KP, FZ, KZ, CLP

Visualization: FY, BMM, ZAA, ZXY, KMF, SNE, PP, WA

Funding acquisition: QGT

Project administration: QGT, FY

Supervision: QGT, YC, CLP

Writing – original draft: FY, QHZ, BMM

Writing – review & editing: QGT, YC, PNM, ZXY, SP

## Data and materials availability

All data are available in the main text or the supplementary materials.

