## Supplementary materials for "Comprehensive Evaluation of Human Donor Liver Viability with Polarization-Sensitive Optical Coherence Tomography"

Feng Yan et al

### METHODS AND MATERIALS

#### Deceased Donor Livers

This study was approved by the University of Oklahoma and the University of Oklahoma Health Sciences Center Institutional Review Board (IRB) (Study number: IRB #12462). There were 20 human deceased donor livers utilized in our experiment that were sourced from the *LifeShare of Oklahoma*. All donor livers used in this study are donation after circulatory death (DCD). The liver samples are delivered to the lab (The University of Oklahoma at Norman) by a professional delivery team, then imaged within 45-90 minutes by PS-OCT, and sectioned within 60-120 minutes after imaging for tissue fixation. None of the livers in this study would be used for transplantation after being studied . The donor's de-identified general profile information was listed in Table S1. The camera images of these 20 donor livers were exhibited in Fig. S8.

#### PS-OCT System and Imaging Protocol

Fig. S9 illustrates the schematic representation of a custom-designed polarization-sensitive optical coherence tomography (PS-OCT) system utilized for imaging *ex vivo* deceased donor livers. The laser source is a broadband superluminescent diode (SLD) with a center wavelength of 1300 nm and a spectral bandwidth of 100 nm. Vertical linearly-polarized light is generated using a polarizer (Pol) and then is directed into polarization-maintaining fibers, subsequently being divided into reference and sample arms through a beam splitter (BS). In the reference arm, the linearly polarized light passes through a zero-order quarter-wave plate (QWP-1) oriented at 22.5 degrees and exits with a 45-degree linear polarization after traversing QWP-1 twice. Within the sample arm, the polarized light passes through a zero-order quarter-wave plate (QWP-2) oriented at 45 degrees, thereby transforming it into circularly polarized light. Upon interaction with the sample, the polarized light undergoes reflection and scattering, resulting in an elliptical polarization state after passing through QWP-2. The recombined polarized light from both arms of the system is then split into the vertical linearly polarized signal and horizontal linearly polarized signal by two polarization-sensitive beam splitters (PBS). Subsequently, these signals are detected and processed by two polarization-sensitive channel sensors (CH-1, CH-2). The detected two orthogonal polarization state lights allow for the analysis of how the tissue alters the polarization state of the light, revealing tissue structure and composition information, particularly regarding the presence of birefringent structures within the tissue. This system provided an axial resolution of 5.5 μm and a lateral resolution of 13 μm in the air with a sensitivity of 105 dB at 48 kHz A-scan rate (*1*). The imaging area at each labeled position was focused on the center of the ink-labeled circle. A field-of-view (FOV) measuring 5 × 5 × 2.6 mm$^3$ (X, Y, Z) with a size of 1000 × 1000 × 1024 pixels in the *X*, *Y*, and *Z* directions was employed, yielding a sampling resolution of 5 × 5 × 2.6 μm$^3$ during liver imaging. The PS-OCT system will generate the Stokes parameters (Q, U, and V) by analyzing the polarization state of light reflected from the sample, which are derived from the intensity measurements of different polarization components of the backscattered light. These parameters are combined to compute the degree of polarization uniformity (DOPU), a metric that quantitatively assesses tissue birefringence by evaluating the consistency of polarization states within a local region (*2*). This process enables the PS-OCT system to noninvasively and efficiently characterize tissue collagen and integrity.

**Histopathological Tissue Processing and Grading**

After PS-OCT imaging, all labeled regions across the entire liver were sectioned and processed for histological staining to compare with corresponding PS-OCT results. At each labeled position on the liver surface, a square area encompassing the central region inside the circle was systematically sampled. The sectioned liver tissue was fixed with 10% formalin, embedded in paraffin, then stained for histological analysis. The biopsy was manipulated and finished by the Tissue Pathology Shared Resource, Stephenson Cancer Center (SCC), and University of Oklahoma Health Sciences Center. The histopathology stains were graded by three board-certified pathologists to generate hepatic viability scores. Each score was finalized through consensus among the three pathologists. In case of dissonant assessments, a consensus workshop was held to establish an agreed-upon final score. To match the imaging penetration depth of PS-OCT (~2 mm) with the pathology examination depth (1.5~2.0 cm), we compared the grading of 30 pathology slides from three donor livers between the full slide thickness (1.5~2.0 cm) and the top 2 mm section, as assessed by the three pathologists. No significant difference in pathological grading between the full slide and the top 2 mm section was found, as shown in Fig. S7. This finding suggests that PS-OCT's imaging depth provides sufficient structural information for viability evaluation.

**Deep-Learning-Based Image Pre-Processing**

To address reflection noise caused by the smooth liver surface and background noise in raw PS-OCT images (red arrows, Fig. S10), we applied deep learning-based image denoising during preprocessing. A total of 1,000 cross-sectional 3D PS-OCT images from each labeled region were preprocessed to enable auto-recognition of steatosis microstructures and texture feature extraction. Pix2Pix, U-Net, Resnet50, and Segment

Anything Model (SAM) model were employed to remove upper-surface reflections and background noise, producing a region-of-interest (ROI) mask. The Resnet50, U-Net, and Pix2Pix models were compiled on Python (Python Org) while the pre-trained Segment Anything Model (SAM) was applied through the Imagesegmenter application on MATLAB (MATLAB Inc.). 4,842 2D PS-OCT intensity images were employed for training models (Resnet50, U-Net, Pix2Pix), 605 images were utilized for validating models (Resnet50, U-Net, Pix2Pix, SAM), and 606 images were used for testing models (Resnet50, U-Net, Pix2Pix, SAM). The accuracy metric evaluates binary pixel accuracy while the intersection over union (IOU) evaluates the accuracy of overlapping regions of the mask generated by the model and the ground truth (manual labeled liver tissue region). The mean square error (MSE) objectively quantifies the magnitude of the errors between the predicted image and the ground truth. The structural similarity index metric assesses quantitative image quality in three structural aspects—luminance, contrast and structure thus overcoming limitations in traditional metrics such as MSE that assume statistical features are spatially stationary (*3*). The peak signal-to-noise-ratio (PSNR) evaluates the quality model segmentation by comparing the ratio between the power of the signal noise and the maximum power of the signal. Higher PSNR scores indicate the higher quality of the masks generated by the model. The Pix2pix model, which includes a modified U-Net generator and a discriminator, outperformed the simple U-Net model, Resnet50 model (*4*), and Segment Anything Model (SAM) (*5*) , as shown in Fig. S10 and Table S2.

The pixel to pixel  (Pix2Pix) is a conditional generative adversarial network (CGAN) that learns to map input images to output images in image-to-image translation tasks (*6*). The Pix2Pix architecture comprises of a U-net-based image generator (*7*) and a convolutional PatchGAN classifier (*6*). The U-Net model consists of an encoder (downsampler) and a decoder (upsampler) with unique skip connections and has the advantage of achieving high segmentation accuracy with fewer training images. The role of the discriminator in the Pix2Pix model is to evaluate each image patch generated by the generator, classifying it as real or synthetic, which helps the generator produce more realistic outputs. In this study, the Pix2Pix model is used to filter noise in PS-OCT images and enhance tissue features by learning the mapping from noisy to clean images through paired training data. This approach enables the model to denoise while simultaneously improving the recognition of liver tissue structures, leveraging its advantage of fewer training parameters and faster compilation times. Specifically, the Pix2Pix generator uses a U-Net architecture with 8 downsampling layers (64-512 filters, LeakyReLU) and 7 upsampling layers (64-512 filters, ReLU, skip connections), with a final output resolution of $256\times256\times3$ and Tanh activation. The discriminator employs a PatchGAN architecture with 3 downsampling layers (64-256 filters, LeakyReLU) and two convolutional layers to classify $30\times30$ patches as real or fake. The model is trained with a batch size of 1, using an adversarial loss (Binary Crossentropy) combined with an L1 loss (weighted by $\lambda=100$), optimized with Adam ($lr=5e\text{-}4$, $\beta1=0.5$).

However, when a filtered image was directly regenerated using the Pix2Pix U-Net model, the original signal of the sample microstructure was altered, introducing errors into subsequent image processing and microstructure recognition. To mitigate this issue, we proposed a mask-assisted segmentation method. In this approach, the Pix2Pix U-Net model generates an ROI mask that is used to segment the denoised ROI from the original image, preserving the original signals (Fig. S11). To evaluate the performance of our method, we compared the results of our mask-assisted filtering approach with those of the direct filtering approach. Six representative regions from the raw image, directly filtered image, and mask-assisted filtered image were selected for comparison. Unlike the direct filtering method, which altered the original signals and introduced errors in microstructures, our mask-assisted filtering method preserved the original PS-OCT intensity signals. These differences were evident in the six selected regions (yellow, cyan, and pink circles within the red, blue, and green frames).

**PS-OCT Imaging for Steatosis Quantification**

The denoised ROI image was flattened to make the liver capsule upper surface remain on the same height in the cross-sectional image. The flattened cross-sectional image set was resliced to the *enface* image. 200 *enface* frames were selected to build a volumetric liver tissue with 500 μm in depth. A Sauvola auto-local threshold algorithm was used to extract the steatosis microstructure from each *enface* frame (Fig. S12) and the 3D steatosis is reconstructed by stacking the 2D steatosis from all selected frames. A raw PS-OCT intensity image was filtered by a 3×3 kernel de-speckle matrix to remove the speckle noise. The denoised intensity image was processed by the Sauvola Auto-Threshold algorithm to segment steatosis microstructures from liver tissues. The recognized steatosis image was inverted and filtered by a 3×3 kernel median filter to remove the recognition errors. The denoised steatosis image was used to quantify the density of steatosis. Fig. S12 shows the overlay of the auto-segmented steatosis with the raw PS-OCT intensity image, which exhibits a high recognition accuracy of our automatic segmentation method.

To compare the segmentation accuracy of hepatic steatosis from PS-OCT intensity images by the Sauvola Auto-Threshold algorithm, three raters were recruited to manually label the hepatic steatosis for the quantification. We provided five representative PS-OCT intensity images with substantial hepatic steatosis. The Sauvola Auto-Threshold algorithm and three raters automatically segmented the steatosis and manually labeled the steatosis for the five images, respectively. Fig. S13A shows the manually labeled steatosis and automatic segmented steatosis from the three raters and the Sauvola Auto-Threshold algorithm. We present the overlay of the manually labeled steatosis from the three raters and found there is a strong match of the labeled steatosis from the three raters. Fig. S13B displays the overlayed steatosis images from the three raters to the corresponding raw PS-OCT intensity images. Dice coefficient was used to evaluate the degree of the agreement between our automatic segmentation method and the rater manual labeling. Fig. S13C is the steatosis density comparison performance between the automatic segmentation and each rater. Our result shows that there is a high match between automatic segmented steatosis and manual labeled steatosis (dice coefficient is in the range of 0.71~0.98). The steatosis density quantification using either our automatic segmentation algorithm or the rater manual labeling shows no significant difference.

**PS-OCT DOPU for Fibrosis Quantification**

The fibrosis signals were extracted from each raw PS-OCT DOPU image by employing five thresholds of 0.5, 0.6, 0.7, 0.8, and 0.9. Five featured fibrosis regions with the DOPU values of 0.5~1.0, 0.6~1.0, 0.7~1.0, 0.8~1.0, and 0.9~1.0 were collected for each DOPU image to quantify the fibrosis through the percentage ratio between the featured fibrosis and the overall hepatic tissue areas. A clinical histopathological score with a range of 0~4 for the fibrosis level of each labeled area was provided by board-certified pathologists as a standard of fibrosis quantification. Each 3D PS-OCT DOPU data has a corresponding histopathologic fibrosis score at each labeled region. To obtain the optimal threshold of obtaining fibrosis levels from PS-OCT DOPU images, linear fitting was performed between each of the five featured percentage ratios and the standard histological score to correlate the pathological score and PS-OCT DOPU data. The threshold with a featured DOPU value corresponding to the highest R-square value in the fitting was selected as the optimal threshold to extract fibrosis signals from hepatic tissues. In this study, 252 labeled areas from 10 donor livers were employed for the correlation between pathology and PS-OCT DOPU images. With the optimal threshold, each percentage ratio of the featured fibrosis area was utilized to present the fibrosis level of the hepatic tissue in each DOPU image. The overall spatial fibrosis score of each labeled area on the donor liver was composed of the average percentage ratio of 1000 2D PS-OCT DOPU images.

**Texture Feature Extraction for Hepatic Inflammation and Necrosis Assessment**

Hepatic inflammation and necrosis, which involve cellular microstructure alterations, are difficult to detect directly using PS-OCT intensity or polarization imaging, especially in the early stages. However, these microstructural changes affected tissue distribution, which can be captured through the pixel distribution characteristics of optical images. Texture features of images describe and quantify spatial and surface properties of objects or regions, such as smoothness, roughness, or coarseness by analyzing the spatial arrangement, size, shape, and spacing of pixels. We applied 27 texture parameters to extract 2,484 texture features from a single 2D PS-OCT intensity frame (multiple features from each texture parameter, as shown in Fig. S14). These parameters are used to extract statistical, structural, model-based, and spatial features of images. The detailed parameters and features have been reported in our previous study (*8*). For each 3D PS-OCT intensity data that is composed of 1,000 2D PS-OCT intensity frames, there are 2,484 × 1,000 2D texture features that were extracted. A 7 × 7 kernel was used to extract 2,484 texture feature values for each pixel within each 2D DOPU image. To obtain the 3D spatial texture feature eigenvalues, the average eigenvalue of 1,000 2D texture feature eigenvalue of each texture feature in the 3D PS-OCT data was calculated (Fig. S14). Thereby 2,484 3D texture feature eigenvalues of each 3D PS-OCT intensity data was generated. To acquire effective texture features for classifying hepatic inflammation and necrosis from PS-OCT intensity images, the unpaired t-student test and random forest learning model were used for the feature screening. 10 donor livers were utilized for effective feature screening by *t*-student test and random forest model. The texture features with the significant difference among groups (Low-Risk *vs* High-Risk and Score 0 *vs* 1 *vs* 2 *vs* 3) based on pathology scores were screened as significant features. A random forest model with a 1.0 training rate and 99% classification accuracy (ROC-AUC=0.99) was employed to screen the effective feature with contributions for classification (weight > 0.0) from significant features. Additionally, the random forest model outputs the weight of each effective feature in classifying inflammation and necrosis. To quantify and visualize the screened effective feature structures from PS-OCT intensity images, a convolutional calculation of each effective feature and the corresponding weight was implemented. The sum of the convolution eigenvalues will be acquired as the composite hyperparameter value.

**Machine Learning Model in Screening Effective Texture Features**

Extracted texture features include effective features (with feature difference) and ineffective features (without feature difference) from PS-OCT intensity images. To screen effective features, we applied machine learning models to classify and verify hepatic tissues with different levels of necrosis and inflammation based on clinical pathology scores. The random forest learning model was trained to classify four scores (0, 1, 2, and 3) and two risk categories (low-risk~0/1 and high-risk~2/3) of necrosis and inflammation from PS-OCT texture feature images based on clinical pathology scores, respectively. A 100% rate of training data was utilized to output the weight of each feature within the classification of hepatic tissues. A threshold of over 0% in the weight of classification contribution was employed to select effective features. The details of effective feature selections by a random forest model are described in Fig. S14. Five supervised learning models (K-Nearest Neighbor (kNN), Gradient Boosting (GB), Decision Tree (DT), Naïve Bayes (NB), and Support Vector Machine (SVM)) were applied to verify the classification performance of the selected effective features with a ratio of 80%: 20% of training versus testing data. To compare the classification performance of our effective feature inputs and other related feature inputs, we applied a k-nearest neighbor (kNN) learning model to classify the hepatic inflammation. 10 donor livers (252 sites) were utilized for comparison. The training accuracy, validation accuracy, and ROC-AUC value were acquired to compare the classification performance from the kNN model with different k numbers (k = 20). Fig. S15A and S15B show that the kNN model with $k \geq 5$ has a significantly higher training and validation accuracy classification with effective feature inputs. Meanwhile, the kNN model has a significantly higher ROC-AUC value under the effective feature inputs, which indicates that our effective feature inputs can improve classification accuracy (Fig. S15C). The parameters that were employed to select the optimal machine learning models were listed in Table S3. The selected optimal parameters for machine learning classifications were provided in Table S4.

### RESULTS

#### Virtual 3D Reconstruction of Liver Score Mapping

To visualize the spatial distribution of liver viability scores across the entire donor liver, we created a virtual three-dimensional (3D) liver model (Fig. S1A, 3DModels.org) and built a 3D reconstruction map of liver viability scores. We recorded the scanning positions on each donor liver and labeled these positions on the virtual 3D model for point labeling (Fig. S1B). This point labeling image was binarized to produce a binary labeling image (Fig. S1C). Next, we extracted edges from the binary labeling image to create an edge labeling image (Fig. S1D). The labeling points were digitized (gray values 0~255) based on the corresponding liver viability scores (PS-OCT or biopsy), resulting in a score labeling image (Fig. S1E). To cover the entire liver surface with scanning points, we expanded the digitized points into digitalized circles using a Gaussian diffusion algorithm. The resulting score diffusion image (Fig. S1F) estimated viability scores for areas not scanned by averaging scores from surrounding digitized circles. We then removed the edges of the liver outlines and the digitized circles to obtain an edge removal diffusion image (Fig. S1G). A liver shape mask was applied to segment the region of interest from this image, creating a shape segmentation image (Fig. S1H). We used a 15×15 kernel Gaussian blur algorithm to filter the shape segmentation image, producing a Gaussian diffusion filtering image (Fig. S1I). Finally, we colorized this image using the Fire color palette to create a Fire color mapping (Fig. S1J), which was overlaid with 25% transparency onto a processed virtual gray 3D model (Fig. S1K), resulting in a virtual color 3D overlap image (Fig. S1L).

#### PS-OCT Imaging of Microstructure and Fibrosis Tissue

PS-OCT image hepatic microstructures and fibrosis by intensity, and polarization images. Fig. S2A shows representative images of hepatic microstructures and fibrosis from intensity, phase retardation, optic axis, DOPU, Stokes-Q, Stokes-U, and Stokes-V. PS-OCT intensity image directly provides the structural imaging of liver tissues such as capsules and blood vessels (Fig. S2B). Stokes parameters (Q, U, V) were used to describe the polarization state of light to characterize the polarization properties of the hepatic tissues. Phase retardation and optic axis qualify the cumulative hepatic fibrosis by using Stokes-Q, Stokes-U, and Stokes-V parameters (Fig. S2C). Additionally, DOPU, calculated by Stokes-Q, -U, and -V parameters, was used to quantify the cumulative hepatic fibrosis.

#### Effective Texture Feature Extraction for Hepatic Inflammation

We screen the effective features for the hepatic inflammation classification and rank effective features based on the corresponding weight. In the specific score classification (i.e., 0 *vs* 1 *vs* 2 *vs* 3), the weight is between 0.0002 and 0.27247 for 310 effective features in 252 imaging sites (Fig. S3A). In the clinical-based thresholding score classification (i.e., Low-risk (0 and 1) *vs* high-risk (2 and 3)), there are 435 effective features screened in 252 imaging sites and the corresponding weight of screened effective features is 0.0005 ~ 0.22512 (Fig. S3B). The feature eigenvalue of all effective features is normalized between 0.0 and 1.0.

**Effective Texture Feature Extraction for Hepatic Necrosis**

We screen the effective features for the hepatic necrosis classification and rank effective features based on the corresponding weight. In the specific score classification (i.e., 0 *vs* 1 *vs* 2 *vs* 3), the weight is between 0.0002 and 0.07515 for 313 effective features in 252 imaging sites (Fig. S5A). In the clinical-based thresholding score classification (i.e., Low-risk (0 and 1) *vs* high-risk (2 and 3)), there are 871 effective features screened in 252 imaging sites and the corresponding weight of screened effective features is 0.0002 ~ 0.16249 (Fig. S5B). The feature eigenvalue of all effective features is normalized between 0.0 and 1.0.

### SUPPLEMENTARY FIGURE LEGENDS

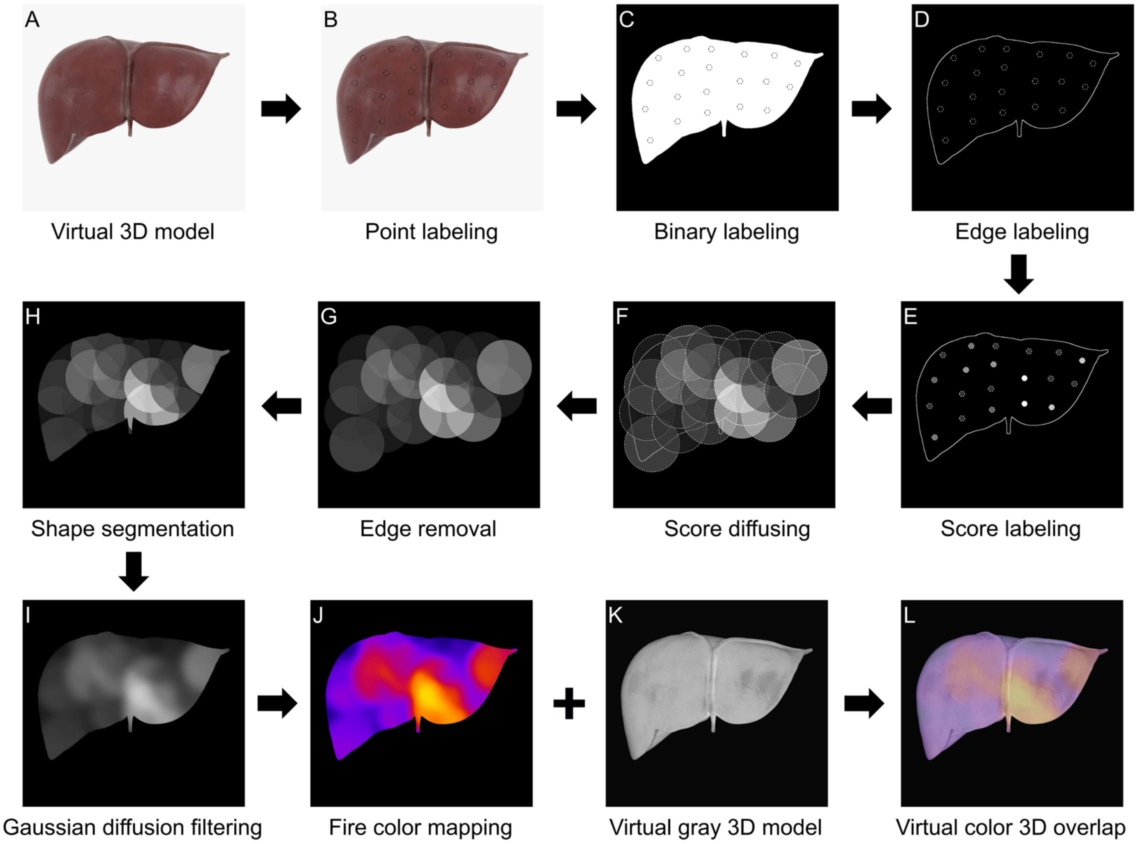


**Fig. S1. The flow diagram of mapping the virtual 3D distribution characters over the entire liver from PS-OCT and biopsy scores.**

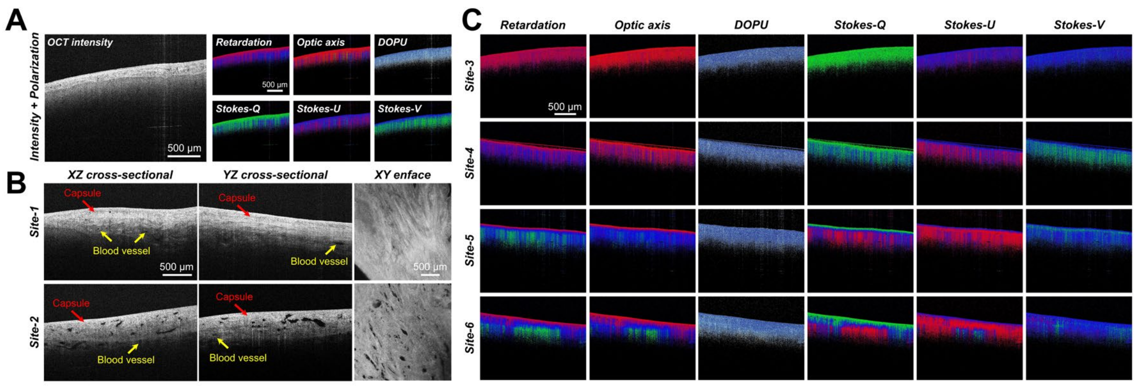


**Fig. S2. Representative image of liver microstructure and fibrosis from PS-OCT at multiple regions.**

**(A)** Representative intensity, phase retardation, optic axis, Stokes-Q, Stokes-U, and Stokes-V images of donor liver tissues from PS-OCT.

**(B)** PS-OCT intensity images of liver microstructures at two different regions from the same donor liver.

**(C)** PS-OCT polarization (phase retardation, optic axis, DOPU) and Stokes (Q, U, V) images of liver fibrosis at four different regions from the same donor liver.

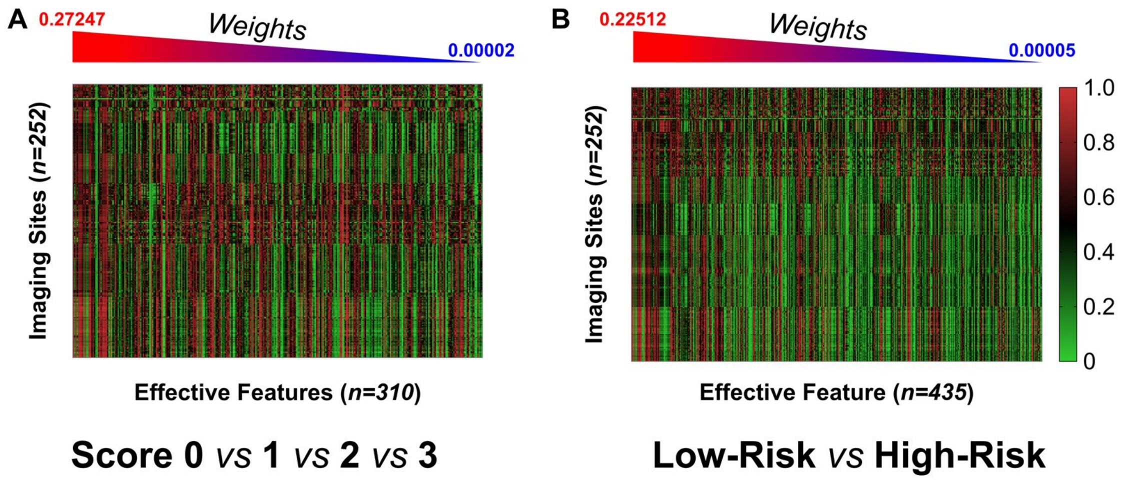


**Fig. S3. Heatmap of screened effective texture feature distributions and corresponding weights for the classification of hepatic inflammation.**

**(A)** The hepatic inflammation classification correlating with specific scores (0, 1, 2, 3).

**(B)** The hepatic inflammation classification correlating with clinical-based thresholding scores (< 2 is Low-Risk, ≥ 2 is High-Risk). N = 12.

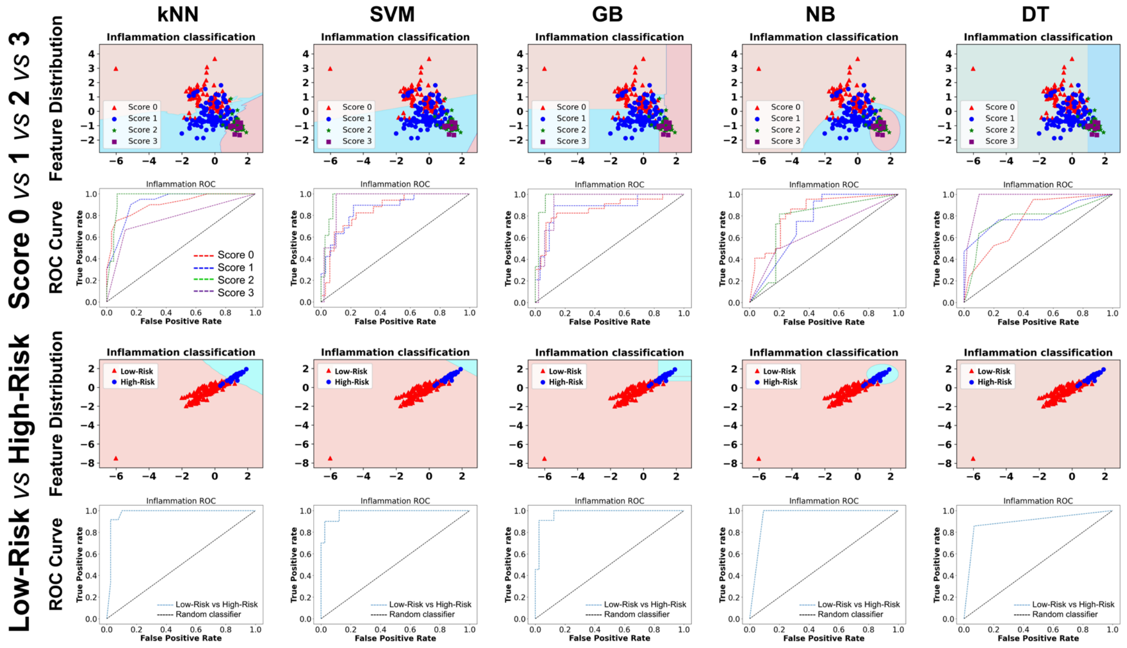

**Fig. S4. The evaluation and performance of machine learning models in hepatic inflammation classification and texture feature distribution based on PS-OCT intensity images.** kNN, k nearest neighbor. SVM, support vector machine. GB, gradient boosting. NB, naïve bayes. DT, decision tree.

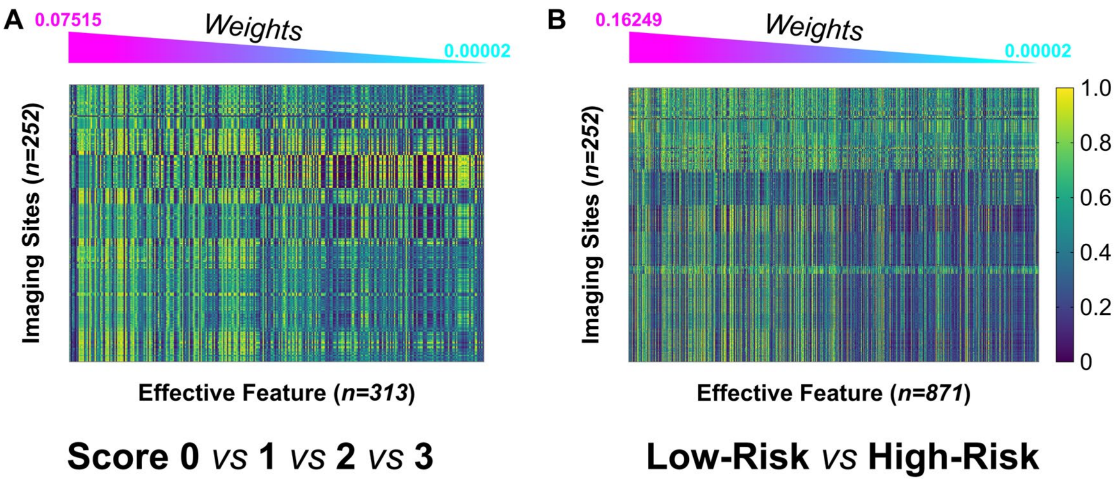


**Fig. S5. Heatmap of screened effective texture feature distributions and corresponding weights for the classification of hepatic necrosis.**

**(A)** The hepatic necrosis classification correlating with specific scores (0, 1, 2, 3).

**(B)** The hepatic necrosis classification correlating with clinical-based thresholding scores (< 2 is Low-Risk, ≥ 2 is High-Risk). N = 12.

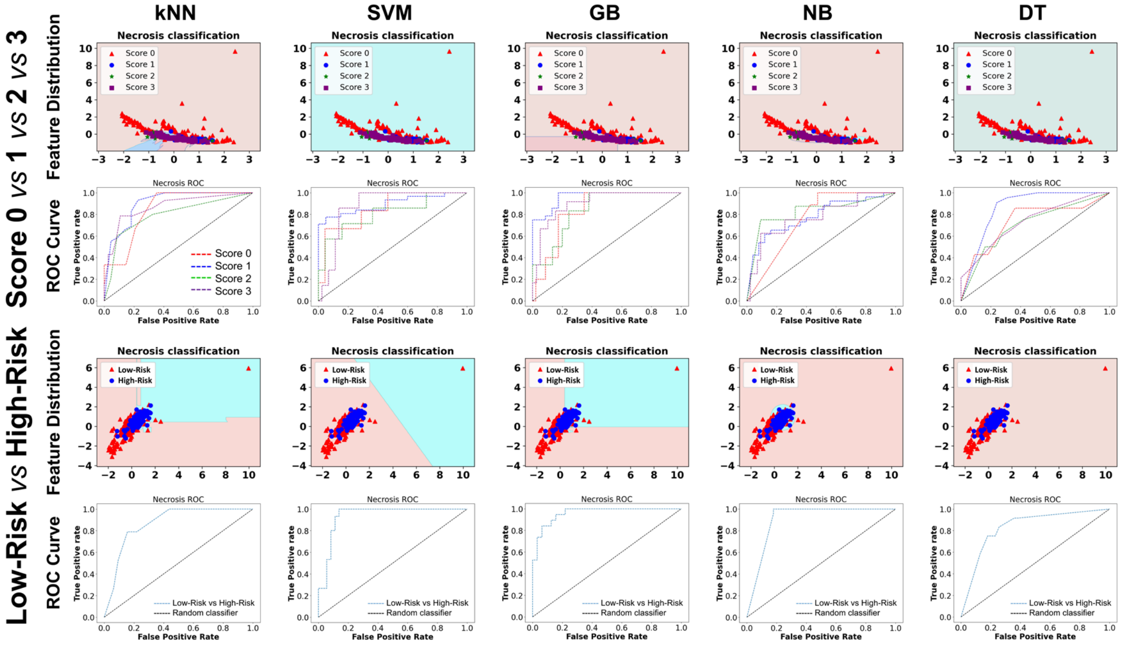

**Fig. S6. The evaluation and performance of machine learning models in hepatic necrosis classification and texture feature distribution based on PS-OCT intensity images.** kNN, k nearest neighbor. SVM, support vector machine. GB, gradient boosting. NB, naïve bayes. DT, decision tree.

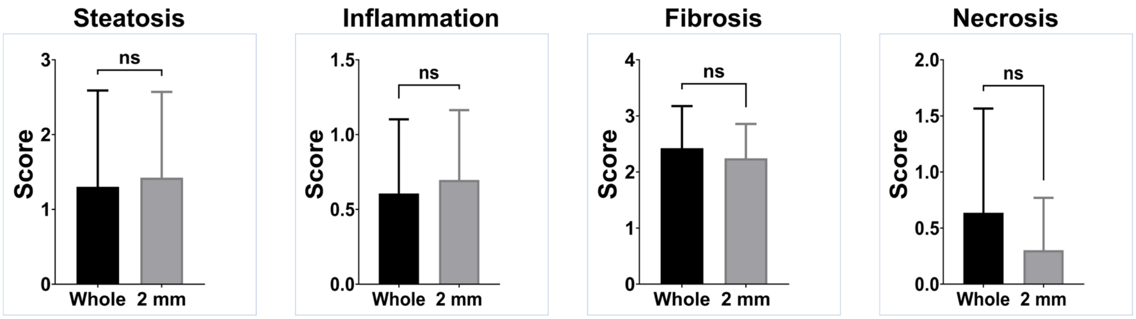


**Fig. S7. The pathological score comparison between the whole slide (1.0~2.0 cm) and the top 2 mm section of slide from three board-certified pathologists.**

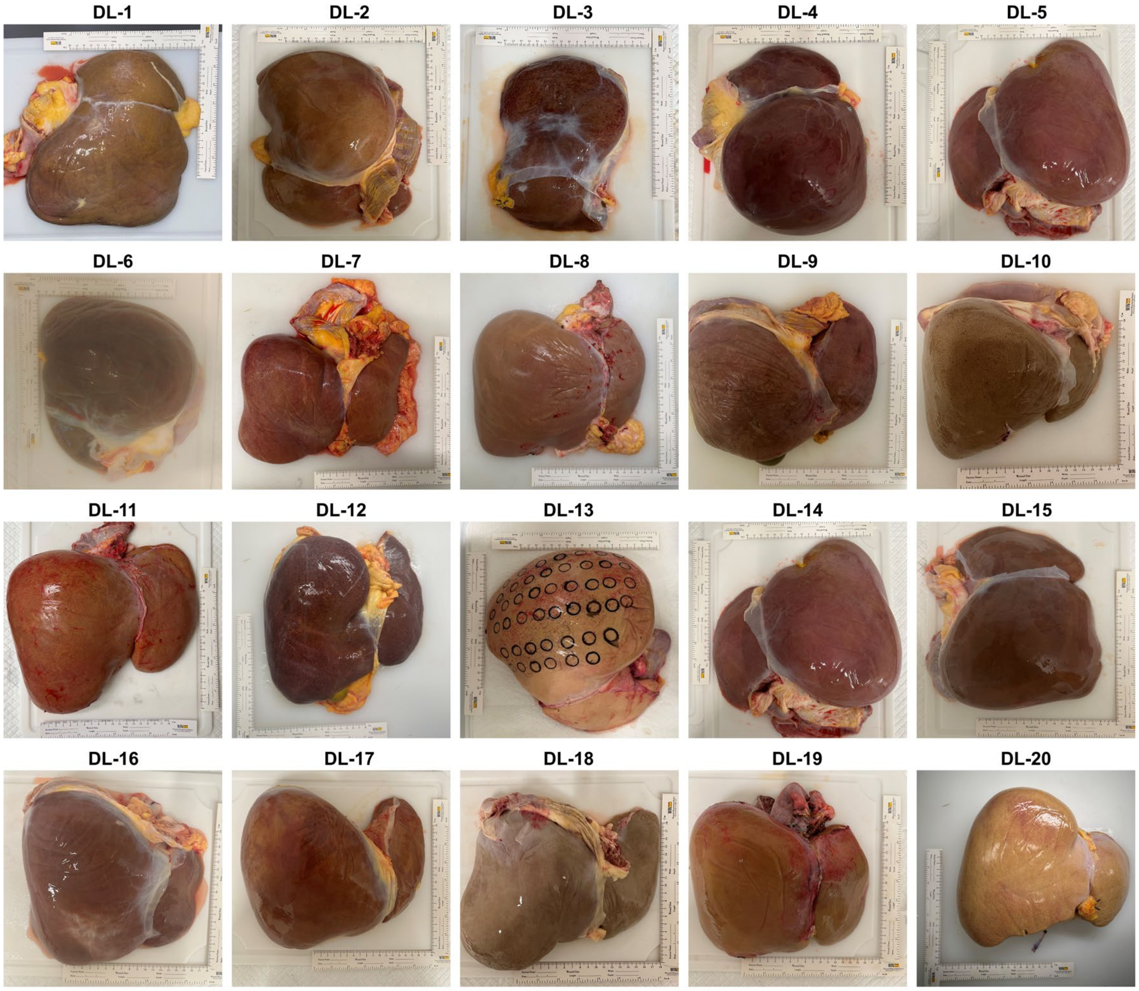


**Fig. S8. Camera images of deceased marginal donor livers (n = 20).** DL, donor liver.

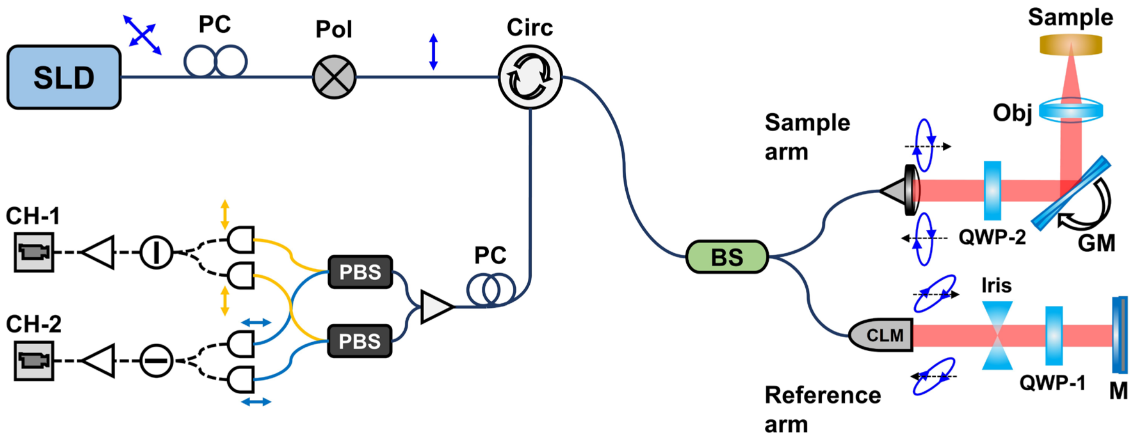


**Fig. S9. The system schematic of polarization-sensitive optical coherence tomography (PS-OCT) for liver imaging.** Broadband Light Source, 1300 nm center wavelength linear-polarized light. PC, polarization controller. Pol, polarizer. Circ, circulator. CLM, fiber-to-free-space collimator. BS, beam splitter. Iris, adjustable iris. QWP-1, quarter-wave plate (orientation ~ 22.5°). QWP-2, quarter-wave plate (orientation ~ 45°). GM, galvanometer. Obj, objective. M, mirror. PBS, polarization-sensitive beam splitter. CH-1, channel-1 sensor. CH-2, channel-2 sensor. Sample Arm of Interferometer, incident circular light – equal light amplitude in both orthogonal polarizations, backscattered and reflected elliptical light – encoded polarization and intensity information.

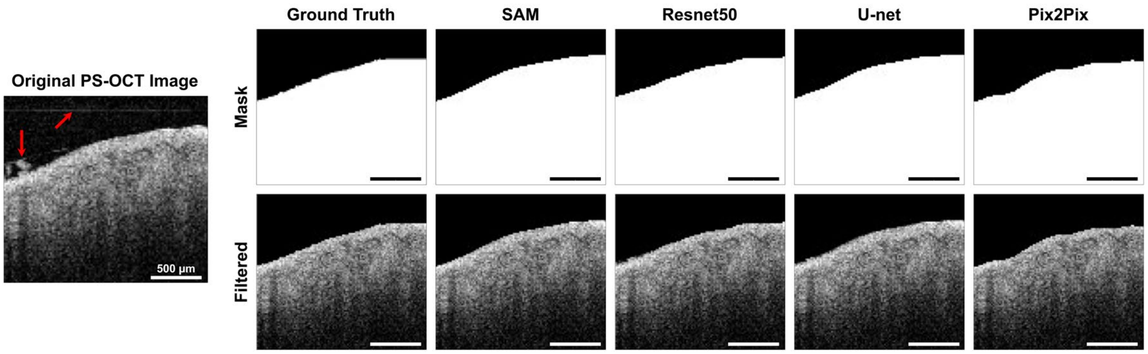


**Fig. S10. Comparison of mask prediction and image filtering for PS-OCT intensity images by Sam, Resnet50, U-net, and Pix2Pix models. SAM, Segment Anything Model.**

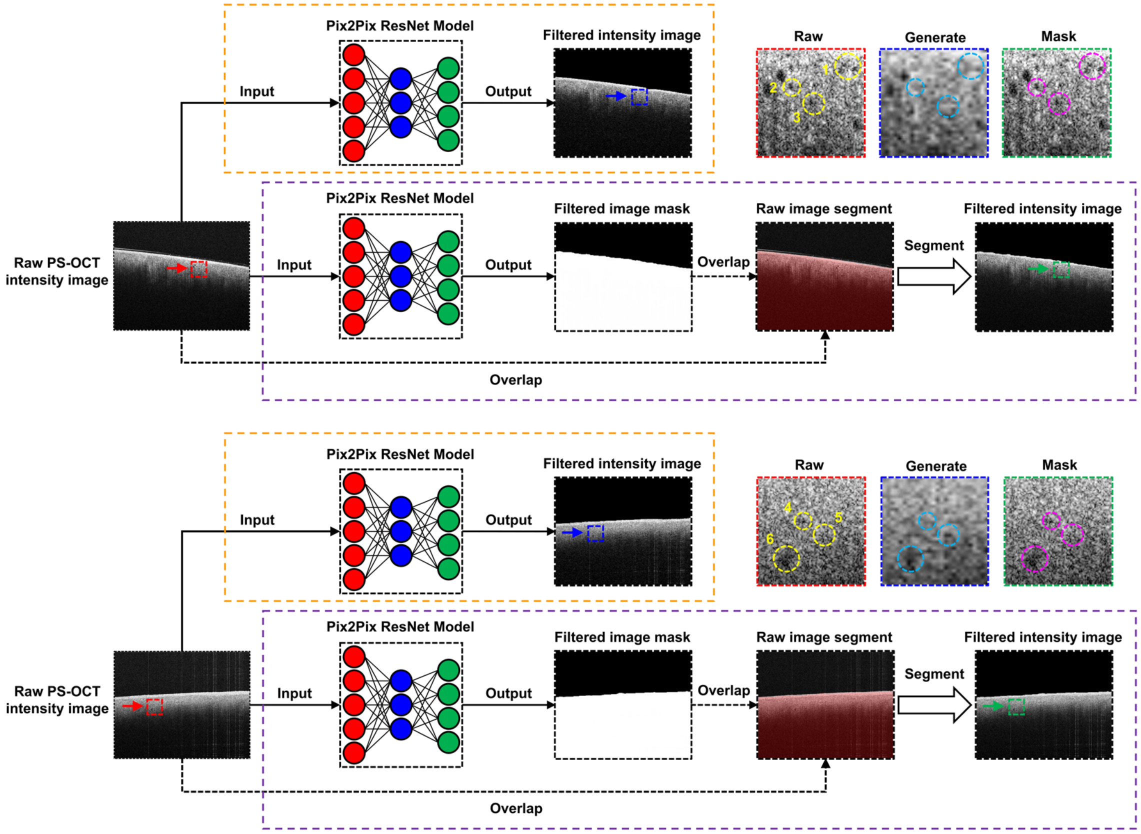


**Fig. S11. Flow chart and diagram of PS-OCT intensity image filtering by a Pix2Pix U-Net model.** Two representative raw PS-OCT intensity images are filtered by the Pix2Pix U-Net model. A direct regenerated filtering image and a mask-assisted filtering image are employed for the image processing. One representative region from the raw PS-OCT intensity image, generated filtering image, and mask-assisted filtering image is selected, respectively.

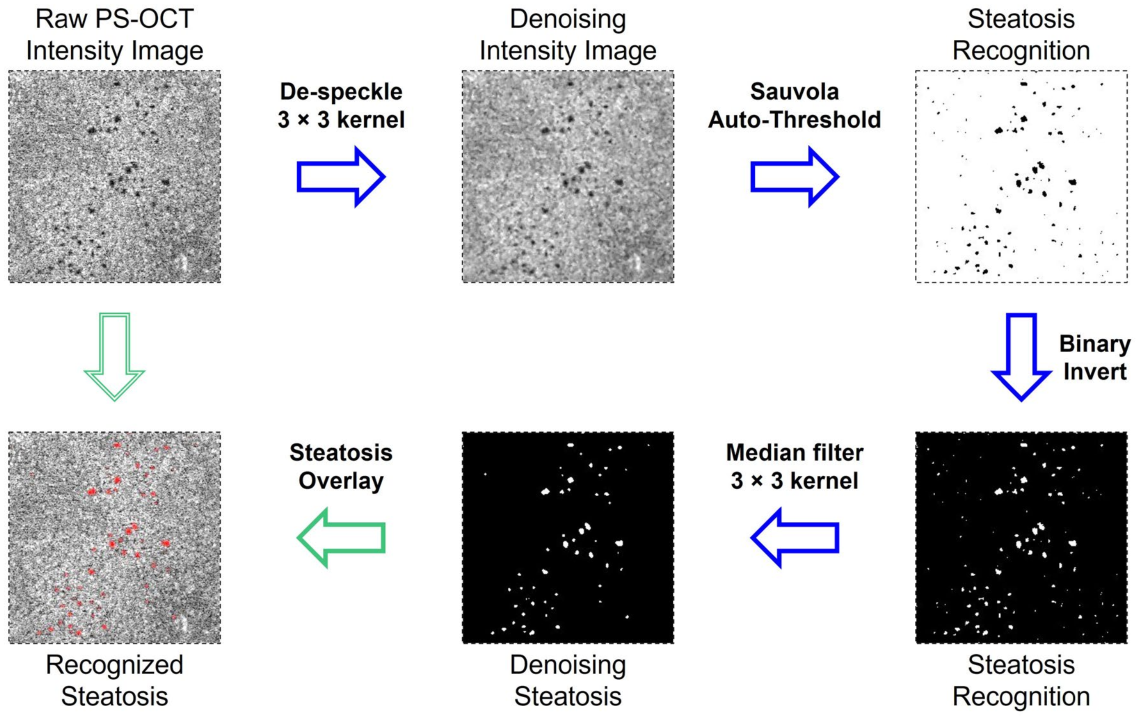


**Fig. S12. Flow diagram of the automatic segmentation of hepatic steatosis from PS-OCT intensity images by automatic threshold segmentation method.**

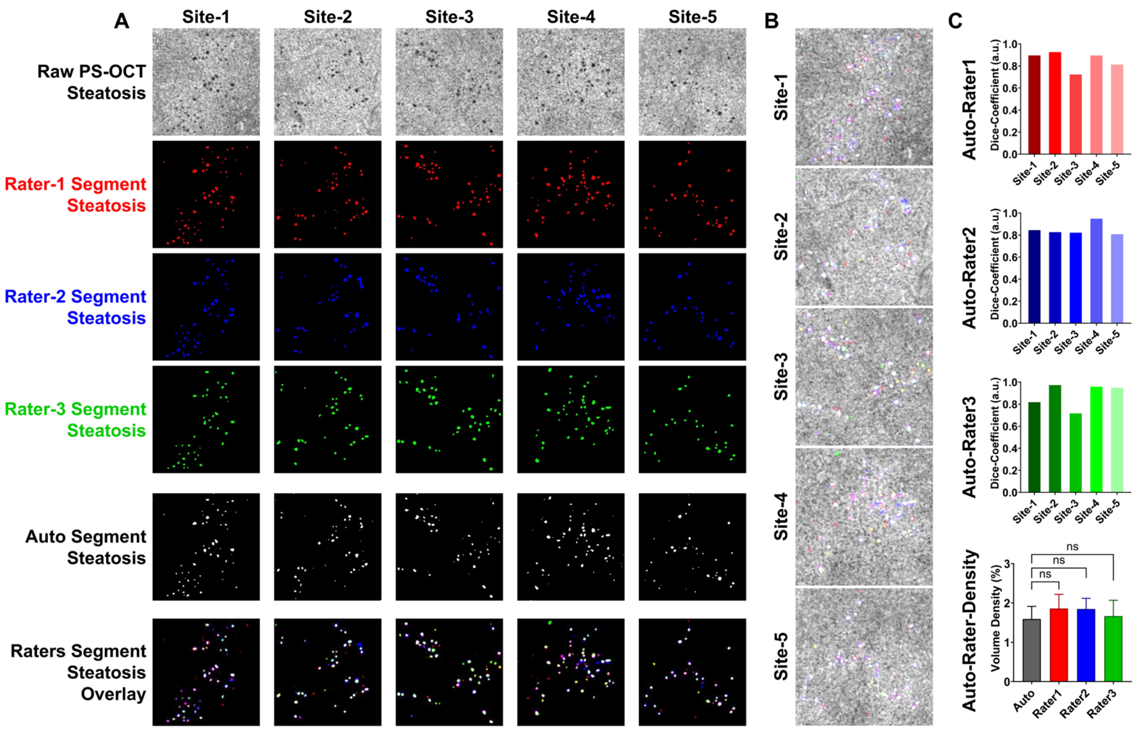


**Fig. S13. Comparison of the segmentation accuracy for the automatic segmentation of hepatic steatosis between the Sauvola Auto-Threshold and manual raters.**

**(A)** Comparison and overlap of recognized hepatic steatosis between automatic segmentation and rater labeling.

**(B)** Overlap between the overlay labeled steatosis of raters and raw PS-OCT intensity images.

**(C)** The comparison of agreement degree between automatic segmentation and rater labeling by dice coefficient and the comparison of steatosis density quantification between automatic segmentation and rater labeling.

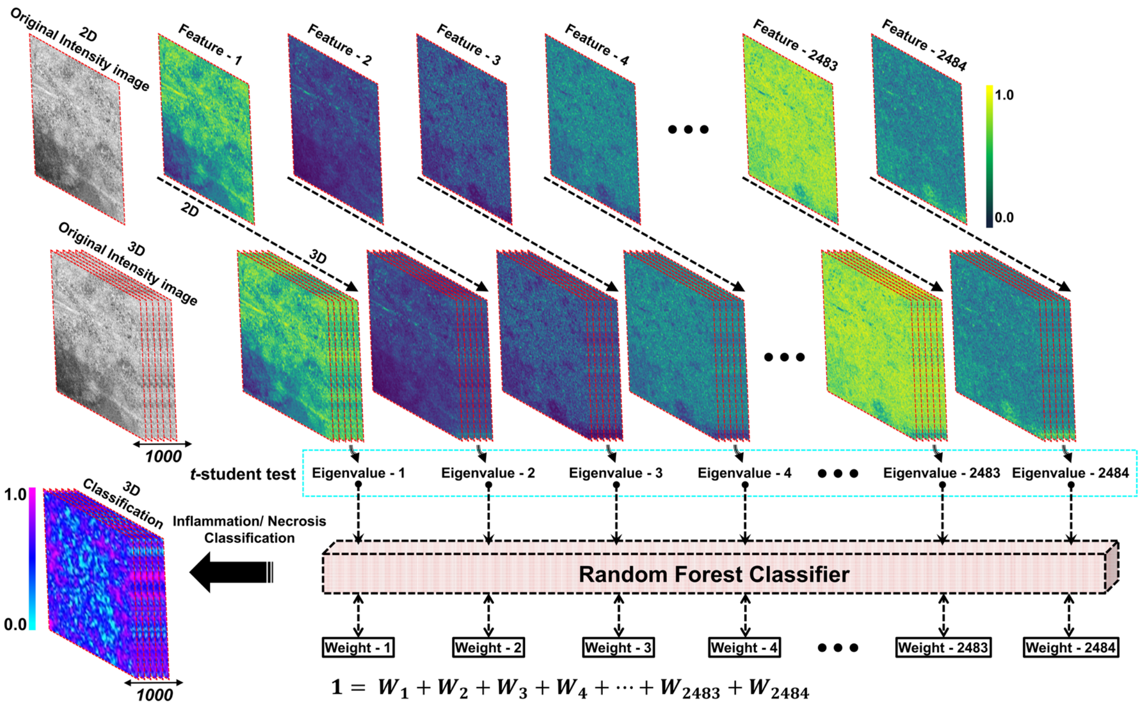


**Fig. S14. Flow chart of the texture feature extraction of PS-OCT intensity images and the screening of effective texture feature.**

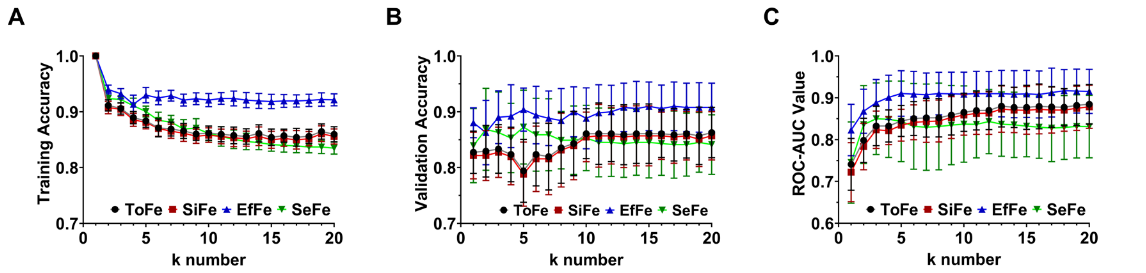


**Fig. S15. Comparison of hepatic inflammation classification accuracy from the k-nearest neighbor (kNN) learning model by different feature inputs.** ToFe, total features (2,484). SiFe, significant features (selected by the unpaired t-student test). EfFe, effective features (screened by the random forest model from significant features). SeFe, selective features (selected by the random forest model from total features).

**(A)** Training accuracy comparison of kNN model with different feature inputs and k numbers.

**(B)** Validation accuracy comparison of kNN models.

**(C)** ROC-AUC comparison of kNN models.

### SUPPLEMENTARY TABLES

**Table S1.** Donor characteristics. BMI, body mass index.

| | Age | Sex | BMI |
|---|---|---|---|
| **DL-1** | 61-70 | M | 28.076 |
| **DL-2** | 51-60 | F | 34.014 |
| **DL-3** | 31-40 | F | 12.458 |
| **DL-4** | 41-50 | M | 29.562 |
| **DL-5** | 51-60 | M | 21.948 |
| **DL-6** | 61-70 | F | 29.2 |
| **DL-7** | 41-50 | F | 38.202 |
| **DL-8** | 31-40 | M | 26.505 |
| **DL-9** | 51-60 | M | 28.839 |
| **DL-10** | 51-60 | F | 23.373 |
| **DL-11** | 21-30 | F | 24.541 |
| **DL-12** | 41-50 | M | 25.728 |
| **DL-13** | 41-50 | M | 24.376 |
| **DL-14** | 41-50 | M | 26.733 |
| **DL-15** | 31-40 | M | 25.794 |
| **DL-16** | 31-40 | F | 22.516 |
| **DL-17** | 31-40 | M | 35.174 |
| **DL-18** | 61-70 | F | 17.032 |
| **DL-19** | 21-30 | M | 21.818 |
| **DL-20** | 51-60 | F | 39.971 |

**Table S2.** Comparison of filtering performance from SAM, Resnet50, U-net, and Pix2Pix models for PS-OCT intensity images. IOU, intersection over union. ROI, region of interest. BGD, background. SSIM, structured similarity index method. PSNR, peak signal to noise ratio. MSE, mean square error.

| Model | Accuracy | IOU (ROI) | IOU (BGD) | IOU (Mean) | SSIM | PSNR | MSE |
|---|---|---|---|---|---|---|---|
| ***SAM*** | 0.9258 | 0.7543 | 0.9034 | 0.8288 | 0.8932 | 12.8160 | 4663.3274 |
| ***Resnet50*** | 0.9517 | 0.8243 | 0.9349 | 0.8796 | 0.9091 | 13.3679 | 3253.8476 |
| ***U-Net*** | 0.9604 | 0.8695 | 0.9495 | 0.9095 | 0.9311 | 18.7757 | 1568.9088 |
| ***Pix2Pix*** | 0.9855 | 0.9384 | 0.9821 | 0.9603 | 0.9514 | 19.4867 | 1008.3915 |

**Table S3.** The range selection of hyper-parameters of machine learning models in the hepatic inflammation and necrosis classification.

| Models | *Parameter* | *Range* |
|---|---|---|
| | *n_neighbor* | 3, 4, …, 19, 20 |
| **kNN** | *weights* | uniform, distance |
| | *algorithm* | auto, ball_tree, kd_tree, brute |

| | | |
|---|---|---|
| | *p* | 1, 2 |
| SVM | *C* | 0.00001, 0.0001, 0.001, 0.01, 0.1 |
| | *kernel* | linear, poly, rbf, sigmoid |
| | *gamma* | 0.00001, 0.0001, 0.001, 0.01, 0.1 |
| GB | *n_estimators* | 100, 200, 300 |
| | *learning_rate* | 0.001, 0.01, 0.1, 0.5, 1.0 |
| | *max_depth* | 3, 4, …, 9, 10 |
| | *min_samples_split* | 2, 3, …, 9, 10 |
| | *min_samples_leaf* | 1, 2, …, 9, 10 |
| | *subsample* | 0.5, 0.6, …, 0.9, 1.0 |
| **NB** | *var_smoothing* | 0.0000000001, 0.000000001, …, 0.01, 0.1 |
| **DT** | *max_depth* | 3, 4, …, 9, 10 |
| | *min_samples_split* | 2, 3, …, 9, 10 |
| | *min_samples_leaf* | 1, 2, …, 9, 10 |
| | *max_features* | none, sqrt, log2 |
| | *max_leaf_nodes* | 10, 20, 30, 40, 50 |
| | *min_impurity_decrease* | 0.0, 0.0001, 0.001, 0.01, 0.1 |

**Table S4.** The selection of optimal hyper-parameters of machine learning models in the hepatic inflammation and necrosis classification.

| **Model** | ***Parameter*** | ***Score 0** vs **1** vs **2** vs **3*** | | ***Low-Risk** vs **High-Risk*** | |
|---|---|---|---|---|---|
| | | *Inflammation* | *Necrosis* | *Inflammation* | *Necrosis* |
| **kNN** | *n_neighbor* | 10 | 10 | 10 | 5 |
| | *weights* | uniform | uniform | uniform | uniform |
| | *algorithm* | auto | auto | auto | auto |
| | *p* | 2 | 2 | 2 | 1 |
| **SVM** | *C* | 0.01 | 0.01 | 0.01 | 0.01 |
| | *kernel* | linear | linear | linear | linear |
| | *gamma* | 0.00001 | 0.00001 | 0.00001 | 0.00001 |
| **GB** | *n_estimators* | 100 | 100 | 100 | 200 |
| | *learning_rate* | 0.01 | 0.01 | 0.01 | 0.01 |
| | *max_depth* | 5 | 3 | 3 | 3 |
| | *min_samples_split* | 5 | 2 | 2 | 10 |
| | *min_samples_leaf* | 10 | 10 | 5 | 10 |
| | *subsample* | 0.8 | 0.8 | 0.8 | 0.8 |
| **NB** | *var_smoothing* | 0.0000001 | 0.0001 | 0.0001 | 0.000000001 |
| **DT** | *max_depth* | 7 | 5 | 10 | 10 |
| | *min_samples_split* | 10 | 2 | 2 | 5 |
| | *min_samples_leaf* | 5 | 5 | 10 | 5 |
| | *max_features* | sqrt | sqrt | sqrt | sqrt |
| | *max_leaf_nodes* | 10 | 10 | 30 | 30 |

| | | | | | |
|---|---|---|---|---|---|
| | *min_impurity_decrease* | 0.01 | 0.01 | 0.1 | 0.01 |
